# Clinically actionable hypertrophic cardiomyopathy genes in South Asian Indians

**DOI:** 10.1101/2025.01.30.25321368

**Authors:** Vinay J Rao, Thiagarajan Sairam, Andiappan Rathinavel, Kurukkanparampil Sreedharan Mohanan, Hisham Ahamed, Jayaprakash Shenthar, Perundurai S Dhandapany

## Abstract

**Background:** Primary hypertrophic cardiomyopathy (HCM) is predominantly a genetic disease causing left ventricular hypertrophy in the absence of other cardiac and systemic metabolic diseases. Currently, limited data exists on the prevalence of clinically actionable gene variants for primary HCM in South Asian Indians (SAI), that is necessary for minimizing disparities in interpreting ancestry-specific variants.

**Objectives:** The ClinGen Hereditary Cardiovascular Disorders (HCVD) Gene Curation Expert Panel categorized HCM-causing genes into five categories according to their clinical relevance: definitive, strong, moderate, limited, and disputed. However, comprehensive studies examining this classification in SAI are lacking.

**Methods:** Whole-exome sequencing was performed for 335 primary SAI-HCM patients, including all known cardiovascular genes and clinically actionable gene categories to determine their allele frequencies.

**Results:** SAI-HCM exomes revealed a total of 194 P/LP and VUS across 26 clinically actionable genes in 119 (35.52%) of the 335 cases. The SAI-HCM cohort exhibited significantly fewer variants in the 12 definitive category genes compared to other global HCM cohorts (17.33% vs. 41.21%, P = 0.0003). For the five strong/moderate genes, no significant difference was observed between the SAI-HCM cohort and other global HCM cohorts (2.59% vs. 2.49%, P = 1). Among the 21 limited and disputed genes, *MYH6* showed a significantly higher prevalence of variants in the SAI-HCM cohort than in the other global HCM cohorts (5.07% vs. 1.67%, P = 0.0408).

**Conclusions:** The clinically actionable gene variants in SAI-HCM cohort differed significantly from other global HCM cohorts, specifically *MYBPC3*, *MYH7*, and *MYH6*.

## Introduction

Hypertrophic cardiomyopathy (HCM) is a diverse genetic heart condition that affects 1 in 500 people globally (1–5). It is defined as thickening of the left ventricle without other cardiac or systemic metabolic disorders. Most HCM cases stem from mutations in genes encoding sarcomere proteins, which have proven useful for accurate clinical diagnosis (6–8). Furthermore, 5-10% of cases are linked to genes not encoding sarcomere proteins (9,10). According to the recent (2024) Clingen curation (https://clinicalgenome.org/), HCM-associated genes were classified into 12 definitive, one strong, four moderate, nine limited, and 12 disputed genes (11) **(gene details, Supplemental Table 1)**. Among the definitive genes, variants in *MYH7* and *MYBPC3* account for the majority of genetically confirmed (7,12–14).

Recent recommendations from the American Heart Association (AHA) and European Society of Cardiology (ESC) emphasize the critical role of genetic testing in managing patients with HCM, identifying at-risk family members early, and improving prognosis (15,16). However, a comprehensive understanding of primary HCM genes remains incomplete owing to the limited diversity of ethnic populations in research. Most genetic research has focused on populations from Europe, Africa-America, the Middle East, East Asia, and SEA (6,17–25). Nevertheless, there remains a gap in the genetic analysis of a substantial Indian cohort, including clinically actionable genes. This limitation hinders the accurate interpretation of genetic variants specific to Indians and other ethnic groups.

Here we present an extensive examination of whole-exome sequences from 335 primary SAI-HCM cases. We compared our findings with those from other global HCM cohorts to determine the difference in the prevalence of ethnic-specific variants.

## Material and methods

### Patient recruitment and selection criteria

A total of 1558 unrelated South Asian (Indian) patients with cardiomyopathy were recruited for this study. These well-characterized individuals came from four medical institutions: Madurai Medical College and Government Rajaji Hospital in Madurai, Government Medical College in Kozhikode, Sri Jayadeva Institute of Cardiovascular Sciences and Research in Bengaluru, and Amrita Institute of Medical Sciences and Research Center in Kochi. Patient consent was obtained in writing, and the study received ethical approval from each participating institution. From this cohort of 1558 participants, 335 cases of primary HCM were identified and selected for further analysis, based on the following criteria and as detailed in **Supplemental Figure 1**. This study was approved by the Institutional Ethical Committee of Institute for Stem Cell Science and Regenerative Medicine, Bangalore (Reference number: inStem/IEC-10/001).

### Inclusion and exclusion criteria for primary HCM patients

- HCM patients, without any secondary or systemic diseases at the time of diagnosis were included
- Cases of HCM linked to conditions such as diabetes, hypertension, hyperlipidemia, coronary artery diseases, valvular diseases, and chronic renal failure, as well as those associated with smoking and alcohol use, were classified as "secondary HCM" and excluded from the study.
- Additionally, the research did not include other forms of cardiomyopathy (such as dilated cardiomyopathy, left ventricular non-compaction, and arrhythmogenic right ventricular dysplasia), syndromic HCM cases, or pediatric HCM patients.

### Population Stratification

The cases were from South India with a confirmed South Asian ancestry, as reported previously using 50 ancestry markers (26).

### Diagnostic criteria for the index individuals

The cases were diagnosed as HCM using a well-established methodology as proposed by the American Society of Echocardiography and as previously described by us (26). A detailed clinical method used for diagnosis is given in **Supplemental Methods**. **Table 1** shows baseline patient characteristics.

**Table 1:** The details of the baseline characteristics of the SAI-HCM patients.

| *Variable* | *SAI-HCM Cohort*<br>*(n=335)* |
| --- | --- |
| ***Demographics*** |  |
| Age, year (mean ± SD) | 44.07±12.77 |
| Male, n (%) | 218 (65.07%) |
| Family history of HCM, n (%) | 100 (29.85%) |
| Competitive sport, n (%) | 1 (0.30%) |
| Chest pain, n (%) | 214 (63.88%) |
| Dyspnea, n (%) | 148 (44.18%) |
| Syncope, n (%) | 194 (57.91%) |
| HbA1c | 4.5±0.6 |
| ***Comorbidities*** |  |
| CAD, n (%) | 0 (0%) |
| Alcohol, n (%) | 0 (0%) |
| Smoking, n (%) | 0 (0%) |
| HTN, n (%) | 0 (0%) |
| Diabetes, n (%) | 0 (0%) |
| Syndromic cases, n (%) | 0 (0%) |
| ***ECG data*** |  |
| Atrial fibrillation, n (%) | 67 (20%) |
| LBBB, n (%) | 14 (4.18%) |
| RBBB, n (%) | 14 (4.18%) |
| AV block, n (%) | 7 (2.09%) |
| T-wave and other abnormalities, n (%) | 251 (74.93%) |
| ***Echocardiography data*** |  |
| LVIDd, mm | 43.63±5.38 |
| LVIDs, mm | 28.15±5.25 |
| LVOTO, n (%) | 117 (34.93%) |
| IVSd, mm | 19.51±3.20 |
| LVEF (%) | 68.56±5.75 |
| ***Septal morphology*** |  |
| Asymmetric, n (%) | 273 (81.49%) |
| Apical, n (%) | 30 (8.96%) |
| Concentric, n (%) | 32 (9.55%) |
| ***Cardiac MRI (n=82)*** |  |
| Presence of LGE, n (%) | 57 (69.51%) |
| ***Intervention/Medication*** |  |
| Myectomy, n (%) | 3 (0.90%) |
| Sudden cardiac death, n (%) | 14 (4.18%) |

ECG: Electrocardiogram; LBBB: Left branch bundle block; RBBB: Right branch bundle block; LVOTO: Left ventricular outflow tract obstruction; LVIDd: Left ventricular internal diameter at diastole; LVIDs: Left ventricular internal diameter at end-systole; IVSd: Interventricular septal thickness at diastole; LVEF: Left ventricular ejection fraction; LGE: Late gadolinium enhancement

### Exome-sequencing and analysis

Genomic DNA was isolated from peripheral blood mononuclear cells of patients using previously published protocols (27). The DNA library was prepared using the SureSelect V5 Enrichment Kit (Agilent). Exome sequencing was performed to generate 100 bp long paired-end reads with a coverage of 100x. We sequenced a total of 20203 genes, which included 39 clinically actionable genes as per the ClinGen HCVD Gene Curation Expert Panel recommendation.

We used an in-house exome analysis pipeline that we previously published (27). Briefly, the raw sequencing reads were trimmed to remove low-quality reads and then aligned to the human reference genome using the Burrows Wheeler aligner (v0.7.17), followed by Picard tools (v 2.9.0) for removal of PCR duplicates, and Genome Analysis Tool Kit (GATK v4.1.9.0) HaplotypeCaller for variant calling. The variants were annotated using ANNOVAR (28). The variants were filtered according to their predicted pathogenicity using SIFT, PolyPhen, and CADD Phred scores (>20). Novel and rare variants (≤ 4*10^-5^; gnomAD v4.1) (6,29) were considered for further analysis. These variants were narrowed down to HCM genes, as outlined in **Supplemental Table 1**. Finally, we classified the obtained variants as pathogenic (P), likely pathogenic (LP), and variants of uncertain significance (VUS) based on ClinVar and American College of Medical Genetics (ACMG) guidelines (29,30). Conflicting VUS and P/LP variants in ClinVar and ACMG were classified as VUS variants **(Supplemental Figure 2)** (29–31).

### Criteria for case excess variant and analysis

Case excess was defined by subtracting the proportion of individuals in controls with a filtered variant from the proportion in the clinical cohort (21,32).

To evaluate case excess, we determined the percentage of surplus or excess variants in the SAI-HCM group (n=335), SA controls (total, n=47177), other global HCM cohorts (n=74–7601) (33–40), and other global controls (n=807162) for further comparison. A detailed breakdown of the examined samples and their characteristics is provided in **Supplemental Table 2**.

### Statistical analysis

Patient baseline characteristics are presented as mean values with standard deviations. Fisher’s exact test was employed to determine the statistical significance of differences in case excess variant numbers between the SAI-HCM cohort and other global HCM cohorts. Statistical significance was defined as a *P*-value< 0.05.

## Results

### Selection of South Asian Indian primary HCM patients

We enrolled a total of 1558 South Asian patients with various types of cardiomyopathies. We excluded 675 cases of secondary HCM, including syndromes, and 548 cases of dilated cardiomyopathy, as outlined in **Supplemental Figure 1**. Finally, we focused solely on the remaining 335 patients with primary HCM.

Among the primary HCM patients (n=335), the average age was 44.07±12.77 years, with males comprising 65.07% of the group. Chest pain (63.88%), syncope (57.91%), and dyspnea (44.18%) were the most frequently reported symptoms. Echocardiographic assessment revealed mean values for left ventricular internal diameter during diastole (LVIDd) and end-systole (LVIDs) of 43.63±5.38 and 28.15±5.25, respectively. The interventricular septum thickness averaged 19.51±3.20, while the mean ejection fraction (EF%) was 68.56±5.75. **Table 1** provides further information on the clinical characteristics of the patients.

### SAI-specific variants in clinically actionable HCM genes

Our analysis revealed 44 pathogenic/likely pathogenic (P/LP) clinically actionable variants in 62 of the 335 primary hypertrophic cardiomyopathy (HCM) cases examined. Of the 44 P/LP variants identified, 24 were novel in the South Asian Indian (SAI) HCM cohort. These novel variants comprised 16 truncating variants found in 20 cases (five in *MYBPC3*, two in *MYH7*, four in *ALPK3*, two in *MYH6*, and one each in *OBSCN*, *RPS6KB1*, and *NEXN*) and nine other variant types, including eight missense, one insertion, and one deletion, observed in 12 cases (Tables 2 and 3, Supplemental Table 3).

**Table 2:** Overall frequencies of case excess variants in clinically actionable definitive and strong gene categories in SAI-HCM.

| Gene | cDNA Variant | Protein Variant | Variant Type | SAI-HCM frequency, n (%) | gnomad global frequency | gnomad SA frequency | ClinVar Classification | ACMG/AMP Classification | Final Classification |
| --- | --- | --- | --- | --- | --- | --- | --- | --- | --- |
| **Definitive** |  |  |  |  |  |  |  |  |  |
| **MYH7** | c.2623_2640del | p.E875_L880del | del | 1 (0.3) | 0 | 0 | NR | LP | LP |
| **MYH7** | c.3985delC | p.L1329Wfs*99 | non | 1 (0.3) | 0 | 0 | NR | LP | LP |
| **MYH7** | c.G4042T | p.E1348X | non | 1 (0.3) | 0 | 0 | NR | LP | LP |
| **MYH7** | c.C1357T | p.R453C | mis | 3 (0.9) | 7.68E-06 | 0 | P | P | P |
| **MYH7** | c.G1988A | p.R663H | mis | 2 (0.6) | 9.92E-06 | 1.10E-05 | P | P | P |
| **MYH7** | c.G2791A | p.E931K | mis | 3 (0.9) | 0 | 0 | P/ LP | LP | LP |
| **MYH7** | c.T2207C | p.I736T | mis | 1 (0.3) | 0 | 0 | P | P | P |
| **MYH7** | c.G2156A | p.R719Q | mis | 1 (0.3) | 0 | 0 | P | P | P |
| **MYH7** | c.G1816A | p.V606M | mis | 2 (0.6) | 4.96E-06 | 0 | P/ LP | P | P |
| **MYH7** | c.C4258T | p.R1420W | mis | 1 (0.3) | 1.30E-05 | 2.19E-05 | P/ LP | P | P |
| **MYH7** | c.C1987T | p.R663C | mis | 1 (0.3) | 2.05E-06 | 0 | P | P | P |
| **MYH7** | c.T743C | p.I248T | mis | 3 (0.9) | 0 | 0 | VUS | LP | VUS |
| **MYH7** | c.G3622A | p.D1208N | mis | 1 (0.3) | 3.61E-06 | 0 | VUS | LP | VUS |
| **MYH7** | c.G4031A | p.R1344Q | mis | 1 (0.3) | 1.18E-05 | 2.20E-05 | VUS | LP | VUS |
| **MYH7** | c.G3779A | p.R1260Q | mis | 2 (0.6) | 1.74E-05 | 3.29E-05 | VUS | VUS | VUS |
| **MYBPC3** | c.1360_1378del | p.V454Wfs*5 | non | 1 (0.3) | 0 | 0 | NR | LP | LP |
| **MYBPC3** | c.3746delG | p.G1249Afs*81 | non | 1 (0.3) | 0 | 0 | NR | LP | LP |
| **MYBPC3** | c.989delC | p.P330Hfs*19 | non | 1 (0.3) | 0 | 0 | NR | LP | LP |
| **MYBPC3** | c.2869dupA | p.T957Nfs*93 | non | 1 (0.3) | 0 | 0 | NR | LP | LP |
| **MYBPC3** | c.C3372A | p.C1124X | non | 5 (1.5) | 1.37E-06 | 0 | P | P | P |
| **MYBPC3** | c.C2526G | p.Y842X | non | 1 (0.3) | 6.842E-07 | 0 | P | P | P |
| **MYBPC3** | c.G1351T | p.E451X | non | 1 (0.3) | 0 | 0 | NR | P | P |
| **MYBPC3** | c.C3811T | p.R1271X | non | 1 (0.3) | 7.46E-06 | 1.10E-05 | P/ LP | P | P |
| **MYBPC3** | c.G1730A | p.W577X | non | 1 (0.3) | 0 | 0 | P/ LP | P | P |
| **MYBPC3** | c.1358delC | p.P453Lfs*12 | non | 1 (0.3) | 0 | 0 | NR | LP | LP |
| **MYBPC3** | c.G2459A | p.R820Q | mis | 1 (0.3) | 2.79E-05 | 1.10E-05 | P/ LP | LP | LP |
| **MYBPC3** | c.G1897A | p.E633K | mis | 2 (0.6) | 0 | 0 | NR | LP | LP |
| **MYBPC3** | c.G1171T | p.D391Y | mis | 2 (0.6) | 3.20E-06 | 2.89E-05 | NR | LP | LP |
| **MYBPC3** | c.T620C | p.L207P | mis | 1 (0.3) | 0 | 0 | NR | LP | LP |
| **MYBPC3** | c.G532A | p.V178M | mis | 1 (0.3) | 1.17E-05 | 2.35E-05 | VUS | VUS | VUS |
| **MYBPC3** | c.G479A | p.R160Q | mis | 1 (0.3) | 2.75E-05 | 0 | VUS | VUS | VUS |
| **TNNT2** | c.411_412insAAA | p.I137_E138insKEE | ins | 1 (0.3) | 0 | 0 | NR | LP | LP |
| **TNNT2** | c.489_490insAAC | p.A163_E164insNR | ins | 1 (0.3) | 0 | 0 | NR | VUS | VUS |
| **TNNT2** | c.C230T | p.P77L | mis | 1 (0.3) | 3.98E-05 | 1.16E-05 | VUS | LP | VUS |
| **TNNT2** | c.G721A | p.E241K | mis | 1 (0.3) | 6.57E-06 | 0 | NR | VUS | VUS |
| **TPM1** | c.226delA | p.K77Rfs*8 | non | 1 (0.3) | 0 | 0 | NR | LP | LP |
| **TPM1** | c.A392G | p.E131G | mis | 1 (0.3) | 0 | 0 | NR | VUS | VUS |
| **MYL2** | c.G64A | p.E22K | mis | 1 (0.3) | 0 | 0 | P/ LP | LP | LP |
| **MYL2** | c.A58G | p.M20V | mis | 2 (0.6) | 6.40E-06 | 1.34E-05 | VUS | VUS | VUS |
| **PRKAG2** | c.1053_1067del | p.L352_E356del | mis | 1 (0.3) | 0 | 0 | NR | VUS | VUS |
| **PRKAG2** | c.C358T | p.R120C | mis | 2 (0.6) | 1.10E-05 | 2.32E-05 | VUS | VUS | VUS |
| **CSRP3** | c.A224T | p.Q75L | mis | 1 (0.3) | 0 | 0 | NR | VUS | VUS |
| **CSRP3** | c.A155C | p.H52P | mis | 1 (0.3) | 1.59E-06 | 0 | NR | LP | LP |
| **FHOD3** | c.1896_1899del | p.E635Gfs*12 | mis | 1 (0.3) | 6.897E-07 | 0 | NR | P (PVS1, PM2) | LP |
| **FHOD3** | c.C4789T | p.R1597X | non | 1 (0.3) | 8.05E-06 | 1.10E-05 | NR | P (PVS1, PM2) | LP |
| **FHOD3** | c.1197_1199del | p.E406del | del | 2 (0.6) | 5.87E-06 | 0 | NR | VUS (PM1, PM2) | VUS |
| **FHOD3** | c.G1040A | p.C347Y | mis | 1 (0.3) | 0 | 0 | NR | VUS (PM1, PM2) | VUS |
| **FHOD3** | c.A2731G | p.T911A | mis | 1 (0.3) | 0 | 0 | NR | VUS (PM1, PM2) | VUS |
| **Strong** |  |  |  |  |  |  |  |  |  |
| **ALPK3** | c.C3772T | p.Q1258X | non | 4 (1.2) | 0 | 0 | NR | P | P |
| **ALPK3** | c.635_636del | p.L213Afs*29 | non | 1 (0.3) | 1.37E-06 | 0 | P/ LP | LP | LP |
| **ALPK3** | c.3575delC | p.R1194Gfs*77 | non | 1 (0.3) | 8.30E-06 | 1.17E-05 | P/ LP | LP | LP |
| **ALPK3** | c.C4768T | p.R1590X | non | 2 (0.6) | 1.30E-05 | 1.10E-05 | VUS | LP | VUS |
| mis = missense; non = nonsense; ins = insertion; del = deletion; NR = not reported |  |  |  |  |  |  |  |  |  |

**Table 3:** Overall frequencies of case excess variants in clinically actionable disputed gene category in SAI-HCM.

| Gene | cDNA Variant | Protein Variant | Variant Type | SAI-HCM frequency, n (%) | gnomad global frequency | gnomad SA frequency | ClinVar Classification | ACMG/AMP Classification | Final Classification |
| --- | --- | --- | --- | --- | --- | --- | --- | --- | --- |
| *Disputed* |  |  |  |  |  |  |  |  |  |
| **MYLK2** | c.111delC | p.P39Qfs*11 | non | 1 (0.003) | 1.24E-06 | 0 | NR | LP | LP |
| **MYH6** | c.5046dupG | p.L1683Afs*3 | non | 1 (0.003) | 0 | 0 | NR | LP | LP |
| **MYH6** | c.C829T | Q277X | non | 1 (0.003) | 2.48E-06 | 3.29E-05 | NR | LP | LP |
| **MYH6** | c.C1720T | Q574X | non | 1 (0.003) | 0 | 0 | NR | LP | LP |
| **MYH6** | c.A5573G | p.E1858G | mis | 10 (0.03) | 1.92E-06 | 0 | NR | VUS | VUS |
| **MYH6** | c.T776C | p.L259P | mis | 1 (0.003) | 0 | 0 | NR | VUS | VUS |
| **MYH6** | c.5125_5151del | p.I1709_L1717del | del | 2 (0.006) | 0 | 0 | NR | VUS | VUS |
| **MYH6** | c.C2623A | p.L875M | mis | 2 (0.006) | 0 | 0 | NR | VUS | VUS |
| **MYH6** | c.T1873C | p.Y625H | mis | 1 (0.003) | 0 | 0 | NR | VUS | VUS |
| **MYH6** | c.G4446C | p.E1482D | mis | 1 (0.003) | 1.24E-06 | 2.20E-05 | NR | VUS | VUS |
| **MYH6** | c.G2037T | p.E679D | mis | 1 (0.003) | 0 | 0 | NR | VUS | VUS |
| **KCNQ1** | c.G676A | p.A226T | mis | 1 (0.003) | 0 | 0 | NR | VUS | VUS |
| **CASQ2** | c.G1132T | p.D378Y | mis | 1 (0.003) | 0 | 0 | NR | VUS | VUS |
| **DSP** | c.C2697G | p.C899W | mis | 1 (0.003) | 0 | 0 | NR | VUS | VUS |
| **DSP** | c.A5963G | p.K1988R | mis | 1 (0.003) | 3.10E-06 | 1.10E-05 | Conflicting | VUS | VUS |
| **DSP** | c.G8031C | p.Q2677H | mis | 1 (0.003) | 0 | 0 | VUS | VUS | VUS |
| **DSP** | c.G3806A | p.R1269Q | mis | 1 (0.003) | 6.20E-06 | 1.10E-05 | Conflicting | VUS | VUS |
| **DSP** | c.A4945G | p.R1649G | mis | 1 (0.003) | 0 | 0 | NR | VUS | VUS |
| **DSP** | c.C5212T | p.R1738X | non | 1 (0.003) | 3.10E-06 | 0 | P | LP | LP |
| **DSP** | c.7683_7685del | p.S2562del | del | 1 (0.003) | 9.91E-06 | 2.20E-05 | VUS | VUS | VUS |
| **MYOM1** | c.G3200A | p.R1067Q | mis | 1 (0.003) | 2.54E-05 | 4.40E-05 | VUS | VUS | VUS |
| **MYOM1** | c.T3331G | p.Y1111D | mis | 1 (0.003) | 1.24E-06 | 2.21E-05 | NR | VUS | VUS |
| **CACNB2** | c.37_39del | p.A16del | del | 1 (0.003) | 3.76E-06 | 1.13E-05 | NR | NR | VUS |
| **CACNB2** | c.G1956T | p.W652C | mis | 1 (0.003) | 0 | 0 | NR | VUS | VUS |
| **ANKRD1** | c.A637C | p.S213R | mis | 1 (0.003) | 1.24E-06 | 0 | NR | VUS | VUS |
| **VCL** | c.G563A | p.R188Q | mis | 1 (0.003) | 6.20E-06 | 0 | NR | VUS | VUS |
| mis = missense; non = nonsense; ins = insertion; del = deletion; NR = not reported |  |  |  |  |  |  |  |  |  |

Among the 24 novel P/LP variants, the *ALPK3* variant p.Q1258X was the most prevalent, occurring in four cases (16.67%). *MYBPC3* had the highest number of novel variants, with seven variants in eight cases (29.17%). Further details of the identified variants are provided in **Table 2**. *MYH7* had the second highest number of novel variants, with six variants in eight cases (25.00%) **(Table 2)**. Two novel variants were found in *MYH6* (8.33%) **(Table 3)**. Single novel variants were identified in *TPM1* (4.17%) **(Table 2)**, *NEXN* (4.17%), and *RPS6KB1* (4.17%) **(Supplemental Table 3)**. Of the novel P/LP variants, the truncating variant *ALPK3* p.Q1258X (16.67%) and missense variant *MYH7* p.E931K (12.5%) were observed most frequently in the SAI-HCM cohort **(Table 2)**.

Furthermore, 59 novel VUSs in 67 cases that were particularly enriched in *OBSCN*, *TTN*, *MYH6*, *RYR2*, and *NEXN* were observed. Among the VUSs, there were no novel *MYBPC3* variants; however, one novel *MYH7* variant was found in three samples, p.I248T (4.48%) **(Table 2)**.

*OBSCN* had the highest number of novel variants (13 variants in 13 samples), followed by *TTN* (12 variants in 14 samples). *MYH6* had five variants in seven samples (10.45%). *RYR2* displayed four novel variants in five samples (7.46%). Three novel variants have been identified in *NEXN* (4.48%) and *DSP* (4.48%). Two novel variants were identified in *FHOD3* (2.99%), *RPS6KB1* (2.99%), *RBM20* (2.99%), and *TMPO* (2.99%) **(Supplemental Table 3)**. In addition, one novel variant in the following genes was discovered: *TPM1* (1.49%), *PRKAG2* (1.49%), *CSRP3* (1.49%), *JPH2* (1.49%), *MYH7* (1.49%)*, TNNT2* (1.49%)*, MYL2* (1.49%)*, CACNB2* (1.49%)*, CASQ2* (1.49%)*, KCNQ1* (1.49%)*, and KLHL24* (1.49%) **(Tables 2 and 3, Supplemental Table 3)**. Eight of the total novel P/LP/VUS variants were truncating (11.94%), two were deletions (2.99%), four were insertions (5.97%), and the remaining 53 were missense variants (79.10%).

Among the novel VUS variants, one missense variant (*MYH7* p.I248T; 4.48%) occurred with the highest frequency in the SAI-HCM cohort **(Table 2)**. In total, among the 119 genotype-positive cases, 35 (10.45%) exhibited compound heterozygosity, while five (1.49%) showed biallelic variants. The remaining 216 cases were classified as idiopathic and lacked known HCM variants.

### Significant differences between SAI-HCM and SA controls in clinically actionable genes

In the SAI-HCM cohort, when compared to the SA controls, we identified significant differences in 7 definitive (*MYH7*, *MYBPC3*, *TNNT2*, *MYL2*, *PRKAG2*, *CSRP3*, *FHOD3*) (19.11% vs. 1.54%, *P* <0.0001), 1 strong (*ALPK3*) (2.39% vs. 0.27%, *P* <0.0001), 1 moderate (*KLHL24*) (0.60% vs. 0.09%, *P* = 0.0386), and 5 limited and disputed genes (*NEXN*, *RBM20*, *OBSCN*, *MYH6*, and *DSP*) (26.28% vs. 6.07%, *P* <0.0001) **(Table 4)**.

**Table 4:**
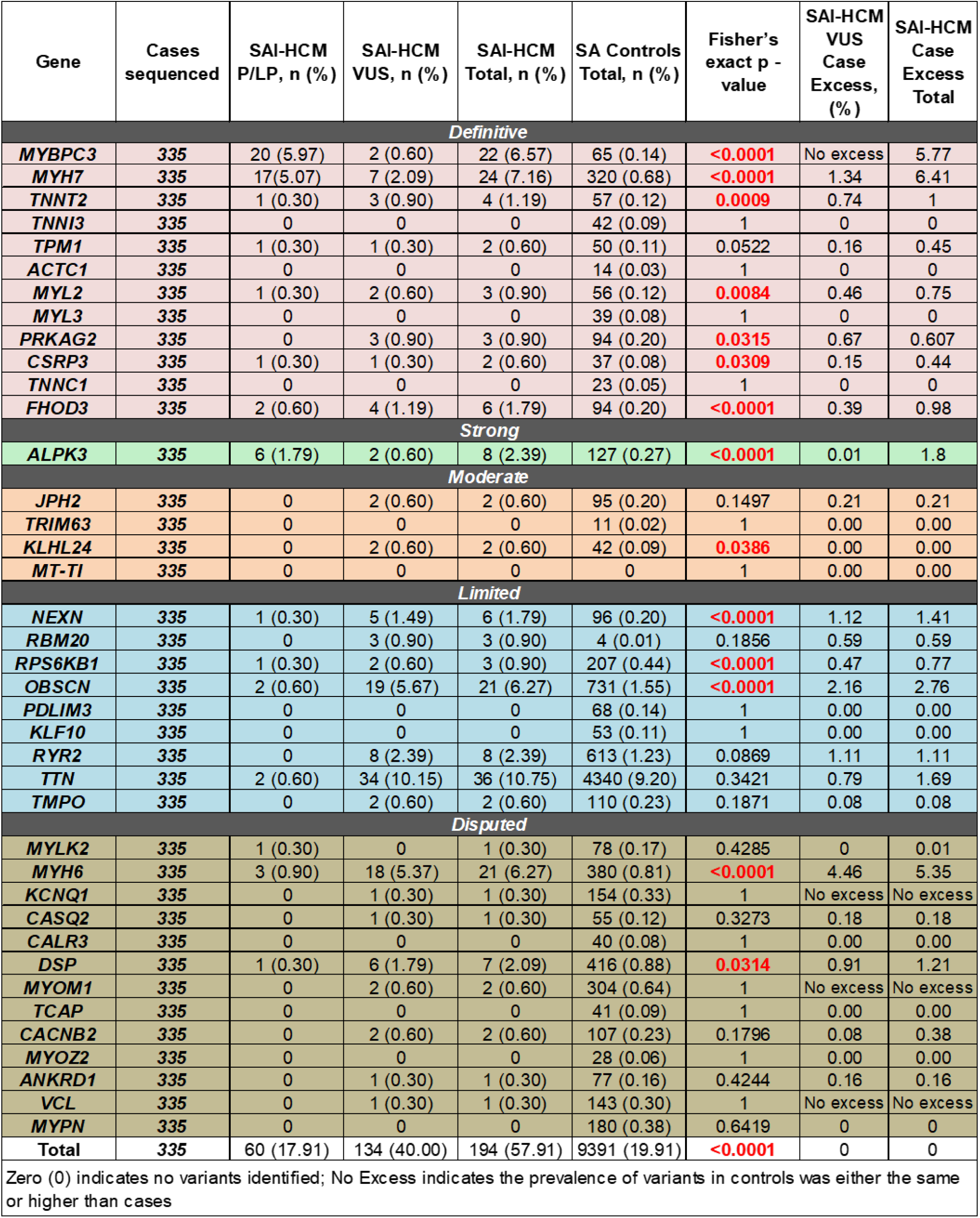
Prevalence of P/LP/VUS variants in SAI-HCM compared to SA controls.

| Gene | Cases sequenced | SAI-HCM P/LP, n (%) | SAI-HCM VUS, n (%) | SAI-HCM Total, n (%) | SA Controls Total, n (%) | Fisher's exact p - value | SAI-HCM VUS Case Excess, (%) | SAI-HCM Case Excess Total |
| --- | --- | --- | --- | --- | --- | --- | --- | --- |
| **Definitive** |  |  |  |  |  |  |  |  |
| *MYBPC3* | 335 | 20 (5.97) | 2 (0.60) | 22 (6.57) | 65 (0.14) | <0.0001 | No excess | 5.77 |
| *MYH7* | 335 | 17 (5.07) | 7 (2.09) | 24 (7.16) | 320 (0.68) | <0.0001 | 1.34 | 6.41 |
| *TNNT2* | 335 | 1 (0.30) | 3 (0.90) | 4 (1.19) | 57 (0.12) | 0.0009 | 0.74 | 1 |
| *TNNI3* | 335 | 0 | 0 | 0 | 42 (0.09) | 1 | 0 | 0 |
| *TPM1* | 335 | 1 (0.30) | 1 (0.30) | 2 (0.60) | 50 (0.11) | 0.0522 | 0.16 | 0.45 |
| *ACTC1* | 335 | 0 | 0 | 0 | 14 (0.03) | 1 | 0 | 0 |
| *MYL2* | 335 | 1 (0.30) | 2 (0.60) | 3 (0.90) | 56 (0.12) | 0.0084 | 0.46 | 0.75 |
| *MYL3* | 335 | 0 | 0 | 0 | 39 (0.08) | 1 | 0 | 0 |
| *PRKAG2* | 335 | 0 | 3 (0.90) | 3 (0.90) | 94 (0.20) | 0.0315 | 0.67 | 0.607 |
| *CSRP3* | 335 | 1 (0.30) | 1 (0.30) | 2 (0.60) | 37 (0.08) | 0.0309 | 0.15 | 0.44 |
| *TNNC1* | 335 | 0 | 0 | 0 | 23 (0.05) | 1 | 0 | 0 |
| *FHOD3* | 335 | 2 (0.60) | 4 (1.19) | 6 (1.79) | 94 (0.20) | <0.0001 | 0.39 | 0.98 |
| **Strong** |  |  |  |  |  |  |  |  |
| *ALPK3* | 335 | 6 (1.79) | 2 (0.60) | 8 (2.39) | 127 (0.27) | <0.0001 | 0.01 | 1.8 |
| **Moderate** |  |  |  |  |  |  |  |  |
| *JPH2* | 335 | 0 | 2 (0.60) | 2 (0.60) | 95 (0.20) | 0.1497 | 0.21 | 0.21 |
| *TRIM63* | 335 | 0 | 0 | 0 | 11 (0.02) | 1 | 0.00 | 0.00 |
| *KLHL24* | 335 | 0 | 2 (0.60) | 2 (0.60) | 42 (0.09) | 0.0386 | 0.00 | 0.00 |
| *MT-TI* | 335 | 0 | 0 | 0 | 0 | 1 | 0.00 | 0.00 |
| **Limited** |  |  |  |  |  |  |  |  |
| *NEXN* | 335 | 1 (0.30) | 5 (1.49) | 6 (1.79) | 96 (0.20) | <0.0001 | 1.12 | 1.41 |
| *RBM20* | 335 | 0 | 3 (0.90) | 3 (0.90) | 4 (0.01) | 0.1856 | 0.59 | 0.59 |
| *RPS6KB1* | 335 | 1 (0.30) | 2 (0.60) | 3 (0.90) | 207 (0.44) | <0.0001 | 0.47 | 0.77 |
| *OBSCN* | 335 | 2 (0.60) | 19 (5.67) | 21 (6.27) | 731 (1.55) | <0.0001 | 2.16 | 2.76 |
| *PDLIM3* | 335 | 0 | 0 | 0 | 68 (0.14) | 1 | 0.00 | 0.00 |
| *KLF10* | 335 | 0 | 0 | 0 | 53 (0.11) | 1 | 0.00 | 0.00 |
| *RYR2* | 335 | 0 | 8 (2.39) | 8 (2.39) | 613 (1.23) | 0.0869 | 1.11 | 1.11 |
| *TTN* | 335 | 2 (0.60) | 34 (10.15) | 36 (10.75) | 4340 (9.20) | 0.3421 | 0.79 | 1.69 |
| *TMPO* | 335 | 0 | 2 (0.60) | 2 (0.60) | 110 (0.23) | 0.1871 | 0.08 | 0.08 |
| **Disputed** |  |  |  |  |  |  |  |  |
| *MYLK2* | 335 | 1 (0.30) | 0 | 1 (0.30) | 78 (0.17) | 0.4285 | 0 | 0.01 |
| *MYH6* | 335 | 3 (0.90) | 18 (5.37) | 21 (6.27) | 380 (0.81) | <0.0001 | 4.46 | 5.35 |
| *KCNQ1* | 335 | 0 | 1 (0.30) | 1 (0.30) | 154 (0.33) | 1 | No excess | No excess |
| *CASQ2* | 335 | 0 | 1 (0.30) | 1 (0.30) | 55 (0.12) | 0.3273 | 0.18 | 0.18 |
| *CALR3* | 335 | 0 | 0 | 0 | 40 (0.08) | 1 | 0.00 | 0.00 |
| *DSP* | 335 | 1 (0.30) | 6 (1.79) | 7 (2.09) | 416 (0.88) | 0.0314 | 0.91 | 1.21 |
| *MYOM1* | 335 | 0 | 2 (0.60) | 2 (0.60) | 304 (0.64) | 1 | No excess | No excess |
| *TCAP* | 335 | 0 | 0 | 0 | 41 (0.09) | 1 | 0.00 | 0.00 |
| *CACNB2* | 335 | 0 | 2 (0.60) | 2 (0.60) | 107 (0.23) | 0.1796 | 0.08 | 0.38 |
| *MYOZ2* | 335 | 0 | 0 | 0 | 28 (0.06) | 1 | 0.00 | 0.00 |
| *ANKRD1* | 335 | 0 | 1 (0.30) | 1 (0.30) | 77 (0.16) | 0.4244 | 0.16 | 0.16 |
| *VCL* | 335 | 0 | 1 (0.30) | 1 (0.30) | 143 (0.30) | 1 | No excess | No excess |
| *MYPN* | 335 | 0 | 0 | 0 | 180 (0.38) | 0.6419 | 0 | 0 |
| **Total** | **335** | **60 (17.91)** | **134 (40.00)** | **194 (57.91)** | **9391 (19.91)** | **<0.0001** | **0** | **0** |

Zero (0) indicates no variants identified; No Excess indicates the prevalence of variants in controls was either the same or higher than cases

Next, we studied case excess, which estimates the excess burden of rare genetic variants in cases compared to controls. An initial case excess study comparing SAI-HCM to SA controls showed a significant increase in P/LP/VUS variants across various gene categories, including definitive (*MYH7*, *MYBPC3*, *TNNT2*, *MYL2*, *PRKAG2*, *CSRP3*, and *FHOD3*), strong (*ALPK3*), moderate (*KLHL24*), limited (*NEXN*, *RBM20*, and *OBSCN*), and disputed (*MYH6* and *DSP*) **(Table 4)**.

### Two definitive HCM genes showed lower variant prevalence in SAI-HCM compared to other global HCM cohorts

*MYBPC3* and *MYH7* are two genes that show the highest frequency of variants in HCM worldwide (7,8,13). However, studies have shown that their case excess in Singaporean (SEA HCM) populations are comparatively lower than those observed in Caucasian (European/American) HCM cohorts. Our findings align with this trend, revealing case excess similar to those of other SEA HCM cohorts (21). Compared to other global HCM cohorts, the SAI-HCM cohort exhibited significantly lower frequencies of P/LP variants in both *MYH7* (5.07% vs. 9.87%, *P* = 0.0024) and *MYBPC3* (5.97% vs. 13.97%, *P* <0.0001). Furthermore, the occurrence of VUSs was also less in *MYH7* (1.18% vs. 3.32%, *P* = 0.0262) and *MYBPC3* (no excess vs. 3.23%, *P* <0.0001) compared to other global HCM cohorts **(Table 5**, **Figure 1)**.

**Figure 1:**
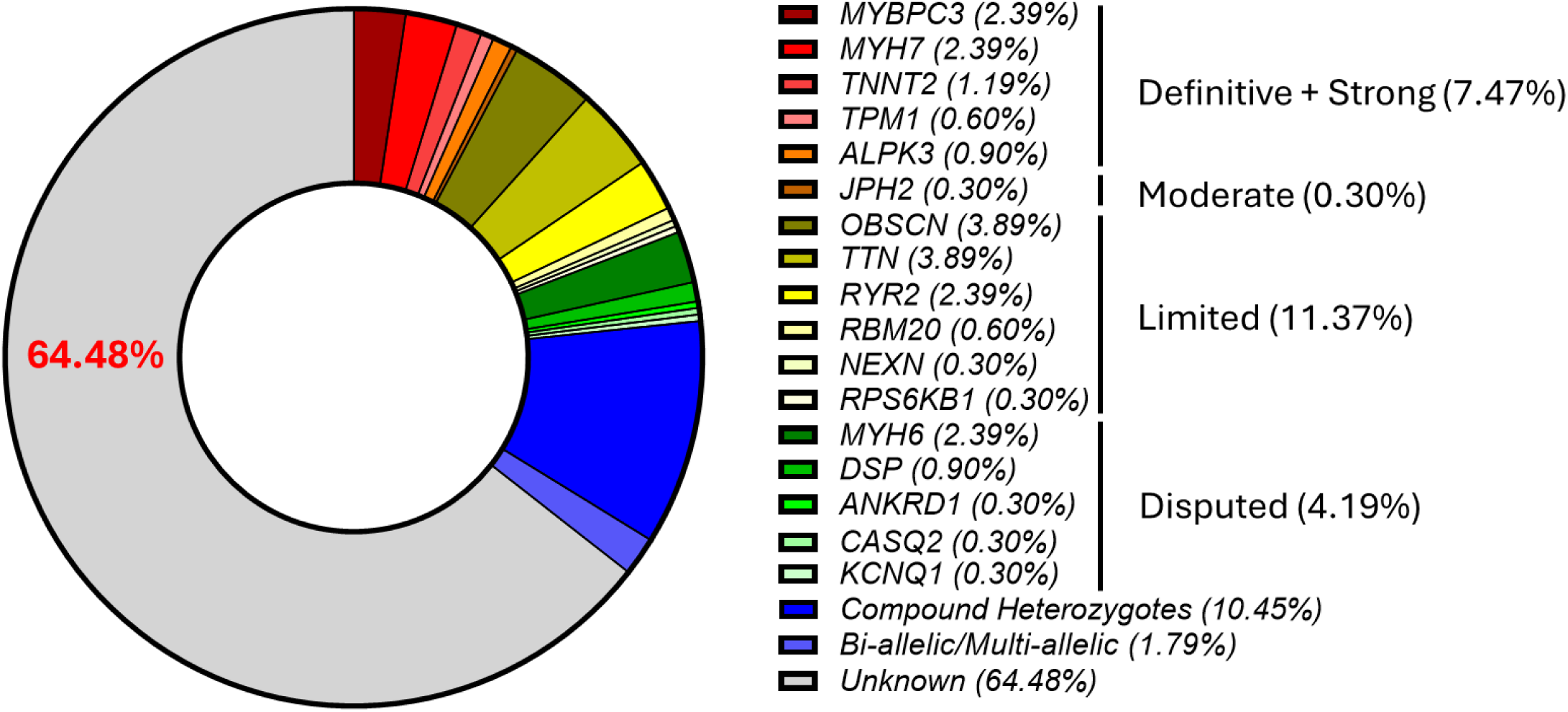
The prevalence of clinically actionable gene variants in SAI-HCM.

**Table 5:** Overall frequencies of case excess variants in clinically actionable genes in SAI-HCM compared to other global HCM cohorts.

| Gene | P/LP (%) |  |  | exVUS (%) |  |  | Case Excess (P/LP/exVUS) (%) |  |  |
| --- | --- | --- | --- | --- | --- | --- | --- | --- | --- |
|  | SAI-HCM<br>(n=335) | Other HCM<br>(n=74 -<br>7601)* | Fisher<br>Exact<br>P Value | SAI-HCM<br>(n=335) | Other HCM<br>(n=74 -<br>7601)* | Fisher<br>Exact<br>P Value | SAI-HCM<br>(n=335) | Other HCM<br>(n=74 -<br>7601)* | Fisher<br>Exact<br>P Value |
| **Definitive** |  |  |  |  |  |  |  |  |  |
| MYH7 | 17 (5.07) | 744 (9.88) | 0.0024 | 4 (1.19) | 250 (3.32) | 0.0262 | 21 (6.27) | 994 (13.19) | < 0.0001 |
| MYBPC3 | 20 (5.97) | 1062 (13.97) | < 0.0001 | No Excess | 246 (3.24) | < 0.0001 | 20 (5.97) | 1308 (17.21) | < 0.0001 |
| TNNI3 | 0 | 105 (1.41) | 0.0241 | 0 | 56 (0.75) | 0.1761 | 0 | 161 (2.16) | 0.0013 |
| TNNT2 | 1 (0.30) | 82 (1.09) | 0.267 | 2 (0.60) | 51 (0.68) | 1 | 3 (0.90) | 133 (1.77) | 0.2879 |
| TPM1 | 1 (0.30) | 33 (0.56) | 1 | 1 (0.30) | 41 (0.70) | 0.7267 | 2 (0.60) | 74 (1.26) | 0.4397 |
| ACTC1 | 0 | 8 (0.14) | 1 | 0 | 17 (0.30) | 0.6207 | 0 | 25 (0.45) | 0.3978 |
| MYL2 | 1 (0.30) | 27 (0.48) | 1 | 2 (0.60) | 28 (0.50) | 0.6854 | 3 (0.90) | 55 (0.98) | 1 |
| MYL3 | 0 | 10 (0.18) | 1 | 0 | 39 (0.70) | 0.1693 | 0 | 49 (0.87) | 0.1132 |
| PRKAG2 | 0 | 12 (0.29) | 1 | 2 (0.60) | 20 (0.48) | 0.6755 | 2 (0.60) | 32 (0.76) | 1 |
| CSRP3 | 1 (0.30) | 8 (0.22) | 0.5525 | 0 | 17 (0.47) | 0.3909 | 1 (0.30) | 25 (0.70) | 0.721 |
| TNNC1 | 0 | 0 | 1 | 0 | 2 (0.04) | 1 | 0 | 2 (0.04) | 1 |
| FHOD3 | 2 (0.60) | 35 (1.10) | 0.5747 | 4 (1.19) | 23 (0.72) | 0.318 | 6 (1.79) | 58 (1.82) | 1 |
| **Strong** |  |  |  |  |  |  |  |  |  |
| ALPK3 | 6 (1.79) | 39 (1.38) | 0.4709 | No Excess | No excess | 1 | 6 (1.79) | 39 (1.38) | 0.4709 |
| **Moderate** |  |  |  |  |  |  |  |  |  |
| JPH2 | 0 | 0 | 1 | 1 (0.24) | 13 (1.06) | 0.3263 | 1 (0.24) | 13 (1.06) | 0.3263 |
| TRIM63 | 0 | 0 | 1 | 0 | 3 (0.05) | 1 | 0 | 3 (0.05) | 1 |
| KLHL24 | 0 | Ins. Nos. | NC | 2 (0.56) | Ins. Nos. | NC | 2 (0.56) | Ins. Nos. | NC |
| MT-TI | 0 | Ins. Nos. | NC | 0 | Ins. Nos. | NC | 0 | Ins. Nos. | NC |
| **Limited** |  |  |  |  |  |  |  |  |  |
| NEXN | 1 (0.30) | 0 | 1 | 4 (1.13) | 0 | 0.1443 | 5 (1.43) | 0 | 0.1443 |
| RPS6KB1 | 1 (0.30) | 2 (1.05) | 0.2978 | 0 | 0 | 1 | 1 (0.30) | 2 (1.05) | 0.2978 |
| RBM20 | 0 | 1 (0.13) | 1 | 2 (0.59) | 12 (1.45) | 0.2528 | 2 (0.59) | 13 (1.58) | 0.2543 |
| OBSCN | 2 (0.60) | 0 | 1 | 5 (1.40) | No excess | 0.5902 | 7 (2.00) | 0 | 0.3596 |
| PDLIM3 | 0 | 0 | 1 | 0 | No excess | 1 | 0 | 0 | 1 |
| KLF10 | 0 | 0 | 1 | 0 | 1 (0.32) | 0.4174 | 0 | 1 (0.32) | 0.4174 |
| RYR2 | 0 | 0 | 1 | 0 | 0 | 1 | 0 | 0 | 1 |
| TTN | 3 (0.90) | 0 | 0.2692 | No Excess | 51 (21.04) | < 0.0001 | 3 (0.9) | 51 (21.04) | < 0.0001 |
| TMPO | 0 | 0 | 1 | No Excess | No Excess | 1 | 0 | 0 | 1 |
| **Disputed** |  |  |  |  |  |  |  |  |  |
| MYLK2 | 0 | 0 | 1 | 0 | 0 | 1 | 0 | 0 | 1 |
| MYH6 | 3 (0.90) | 0 | 0.2692 | 14 (4.18) | 4 (1.67) | 0.0957 | 17 (5.07) | 4 (1.67) | 0.0408 |
| KCNQ1 | 0 | 0 | 1 | No Excess | 5 (1.94) | 0.0124 | 0 | 6 (2.5) | 0.0231 |
| CASQ2 | 0 | 0 | 1 | 1 (0.25) | 0 | 1 | 1 (0.25) | 1 (0.42) | 1 |
| CALR3 | 0 | 0 | 1 | 0 | 2 (0.65) | 0.1738 | 0 | 2 (0.65) | 0.1738 |
| DSP | 1 (0.30) | 0 | 0.2509 | No Excess | No Excess | 1 | 1 (0.30) | 0 | 0.2509 |
| MYOM1 | 0 | 0 | 1 | No Excess | No Excess | 1 | 0 | 0 | 1 |
| TCAP | 0 | 0 | 1 | 0 | 0 | 1 | 0 | 0 | 1 |
| CACNB2 | 1 (0.30) | Ins. Nos. | NC | No Excess | Ins. Nos. | NC | 1 (0.30) | Ins. Nos. | NC |
| MYOZ2 | 0 | 0 | 1 | 0 | 0 | 1 | 0 | 0 | 1 |
| ANKRD1 | 0 | Ins. Nos. | NC | 0 | Ins. Nos. | NC | 0 | Ins. Nos. | NC |
| VCL | 0 | 0 | 1 | No Excess | No Excess | 1 | 0 | 0 | 1 |
| MYPN | 0 | 0 | 1 | 0 | 3 (1.25) | 0.0722 | 0 | 3 (1.25) | 0.0722 |
| **Total** | **61 (18.21)** | **2168 (28.52)** | **< 0.0001** | **44 (13.13)** | **881 (11.59)** | **0.3843** | **105 (31.34)** | **3066 (40.34)** | **< 0.0001** |

NC = Not comparable; Ins. Nos. = Insignificant numbers; cannot be used for analysis

### Three definitive genes are underrepresented in the SAI-HCM cohort

Notably, three definitive genes (*ACTC1*, *MYL3*, and *TNNC1*), which represented 1.36% of the variants in other global HCM cohorts, showed no P/LP or VUS variants in the SAI-HCM cohort **(Table 5**, **Figure 1)**. *TNNI3* variants (another definitive gene) are significantly more prevalent in the SEA HCM cohort (7.7%) than in the European HCM cohort (2.00%) (13,21) and other Indian cohorts (3%) (41). However, no variants were observed in *TNNI3* in this study.

### *MYH6* (a disputed gene) variants is significantly increased in SAI-HCM compared to other global HCM cohorts

In the limited gene category, *NEXN* was found to have an insignificant case excess in the SAI-HCM cohort than in other global HCM cohorts (1.42% vs. 0.05%, *P* = 0.0786). In addition, *RPS6KB1*, *OBSCN*, *RBM20*, *PDLIM3*, *KLF10*, *RYR2*, and *TMPO*, and *TTN* showed similar or less frequency in the SAI-HCM and other global HCM cohorts **(Table 5)**.

Among the disputed genes, including *MYLK2*, *CASQ2*, *CALR3*, *DSP*, *MYOM1*, *TCAP*, *CACNB2*, *MYOZ2*, *ANKRD1*, and *VCL*, no significant P/LP/VUS variants were observed. Interestingly, *KCNQ1* showed significantly less case excess in the SAI-HCM group compared to other global HCM cohorts (0.30% vs. 2.50%, *P* = 0.0231) **(Table 5)**. Conversely, *MYH6* demonstrated an increased case excess in the SAI-HCM cohort relative to other global HCM populations (5.07% vs. 1.67%, *P* = 0.0408) **(Table 5)**. Notably, half of the *MYH6* variants were identified as compound heterozygotes (47.62%) **(Table 5)**.

### Compound heterozygosity in the SAI-HCM cohort

Out of 335 cases of HCM, 119 were genotype-positive for clinically actionable genes. Within this group, 35 samples exhibited compound heterozygosity, representing 10.45% of genotype-positive individuals **(Supplemental Table 4)**. This proportion aligns with the 3-19% range reported across various HCM cohorts in a recent meta-analysis study (42). We also identified five instances of biallelic variants and one case of multiallelic variants **(Supplemental Table 4)**.

### Case excess variants of SAI-HCM are distinct from other global HCM cohorts

When comparing SAI-HCM to other global HCM cohorts, we observed lower percentages of case-excess variants in definitive genes, such as *MYH7* (6.26%) and *MYBPC3* (5.56%) **(Table 5**, **Figure 1, Supplemental Table 5)**. For the remaining definitive HCM genes (*ALPK3*, *TNNT2*, *TPM1*, *MYL2*, and *CSRP3*), P/LP variants constituted 3.54% of our cohort, compared to 4.13% globally **(Table 5**, **Figure 1, Supplemental Table 5)**. The SAI-HCM cohort demonstrated a slight, statistically non-significant increase in *ALPK3* frequency when compared to other global HCM cohorts (1.42% vs. 0.6%, *P* = 0.0797) **(Table 5, Supplemental Table 5)**.

The SAI-HCM cohort showed no P/LP variants in moderate genes. Within the limited category of HCM-associated genes, we detected a total burden of 2.06% P/LP variants, which included *NEXN* (0.29%) and *RPS6KB1* (0.29%) **(Table 5**, **Figure 1, Supplemental Table 5)**.

Among the disputed category, most genes had no significant differences, while only *KCNQ1* displayed fewer variants in the SAI-HCM cohort compared to other global HCM cohorts. In contrast, the case excess of *MYH6* variants was found to have a significant increase in the SAI-HCM cohort than in the other global HCM cohorts **(Table 5, Supplemental Table 5)**.

In the disputed category, most genes exhibited similar frequencies in SAI-HCM compared to other global HCM cohorts. However, *KCNQ1* was an exception, showing fewer variants in the SAI-HCM cohort compared to other global HCM cohorts (0.30% vs. 2.50%, *P* = 0.0231).

Conversely, *MYH6* variants demonstrated a significant increase in the SAI-HCM cohort relative to other global HCM cohorts, indicating case excess **(Table 5, Supplemental Table 5)**.

In the SAI-HCM cohort, we identified 36 case excess VUSs, accounting for 10.55% of the total. These were distributed across categories, with 3.3% in the definitive group, 0.24% in the moderate group, and 7% in the limited group **(Table 5, Supplemental Table 5)**.

## Discussion

To our knowledge, this study constitutes a relatively large SAI primary HCM cohort (n=335), employing rigorous selection criteria to detect clinically relevant gene variants. While previous investigations have attempted to elucidate the genetic underpinnings of HCM in SAI, they were restricted by factors such as case reports and utilization of targeted or clinical exome panels with limited sample sizes (43–47). Also, an important pitfall in such investigations is the inclusion different types of cardiomyopathies, including secondary and syndromic HCM, to determine the frequency of clinically actionable gene variants for primary HCM. Such inclusions can lead to inaccurate attribution of specific pathogenic variants to primary HCM. To address this issue, our research focused exclusively on primary HCM cases.

Various studies in other global HCM cohorts have also relied solely on clinical exome panels (21,35–38). Recent investigations have highlighted the contrast between targeted sequencing and exome sequencing, with the former identifying variants in restricted genes, thereby excluding other potentially disease-causing or modifier genes (48). In contrast, our study has focused on whole-exome sequencing of primary HCM and analyzing all the known HCM genes till date.

This approach yielded 61 P/LP variants and 67 excess VUSs, including 24 novel P/LP and 59 novel VUSs in 119 SAI-HCM patients (total n=335), collectively accounting for 35.52% of the HCM cases **(Table 5**, **Figure 1)**. We did not observe any P/LP/VUS variants in the remaining 13 clinically actionable HCM genes and other inheritable cardiac disease genes (n=166) in the SAI-HCM cohort, except one mono-allelic variant in *LZTR1* (p.E331X) **(Supplemental Table 6)**.

Notably, mono-allelic *LZTR1* variants were observed in multiple healthy individuals, and mostly bi-allelic variants cause Noonan syndrome as reported by Johnston *et al* (49).

Among the 119 SAI-HCM genotype-positive patients, we identified thirty-five cases with compound heterozygotes (10.45%) **(Supplemental Table 4)**. These data suggest that SAI bear a similar burden to individuals from SEA (21) but different from European, American, and Middle Eastern cohorts. However, the SAI-HCM cohort showed a higher case excess in *MYH6* compared to SEA and other global HCM cohorts. Although disputed, various genetic and functional studies support the role of *MYH6* in sarcomeric disorganization and HCM (50–54).

Multiple investigations focusing on clinical exomes (not whole exomes) in Indian HCM patients, albeit with fewer cases, have shown reduced frequencies of variants in *MYBPC3*, *MYH7*, *TPM1*, and *TNNT2* genes (43,55–57) (with the exception of *TNNI3* (41)). Consistent with these findings, our current study on the SAI-HCM cohort also identified similar or fewer clinically relevant variants in *MYH7*, *MYBPC3*, *TNNT2*, and *TNNI3*. Notably, these gene variants are highly common in other global HCM cohorts.

A comprehensive meta-analysis, encompassing 455 research studies with a total of 20,808 HCM samples from diverse ancestries, revealed that sarcomeric gene variants (including *MYBPC3*, *MYH7*, *TNNT2*, *TNNI3*, *MYL2*, *MYL3*, *TPM1*, *ACTC1*) occurred in 19-53% of cases, with an average prevalence of 34% (58). Our findings in the SAI-HCM cohort demonstrated a comparable prevalence rate.

Genetic testing for pathogenic variants in patients with HCM does not consistently lead to conclusive interpretation and diagnosis because multiple populations are underrepresented in research studies. For example, the recent ClinGen curation has downgraded *MYH6* to disputed from limited category (11). However, our findings revealed a high prevalence of *MYH6* in the SAI-HCM cohort, which differs from other global HCM cohorts. These findings underscore the need to examine clinically relevant HCM genes across diverse ethnic backgrounds and in underrepresented populations. Such comprehensive testing will improve the guidelines for interpreting pathogenic variants and genetic-based diagnostic procedures.

## Conclusion

Taken together, in our case excess study, we identified that 7 definitive (*MYH7*, *MYBPC3*, *TNNT2*, *MYL2*, *PRKAG2*, *CSRP3*, *FHOD3*), 1 strong (*ALPK3*), 1 moderate (*KLHL24*), and 5 limited and disputed genes (*NEXN*, *RBM20*, *OBSCN*, *MYH6*, and *DSP*) were significantly correlated with HCM in SAI-HCM patients as compared to SA controls. However, their prevalence when compared individually to other global HCM cohorts was either similar (*TNNT2*, *CSRP3*, *FHOD3*, *ALPK3*, and *KLHL24*) or lower (*MYH7*, *MYBPC3*, *TNNI3*, *TTN*, and *KCNQ1*).

Only *MYH6* displayed a higher prevalence of variants compared to other global HCM cohorts. These data suggest that SAI patients exhibit overlapping and distinct HCM allele frequencies compared to other global HCM cohorts.

Our study identified clinically significant genes in only 35.52% (n=119) of the 335 SAI-HCM patients. The genetic cause remains unknown for 64.48% (n=216) of Indian HCM patients (Figure 1). This study represents the first comprehensive exome analysis of Indian patients with primary HCM, highlighting the importance of understanding the ethnicity-specific genetic architecture of HCM for precise diagnosis, risk evaluation, and treatment strategies.

## Limitations

Despite employing a comparatively substantial sample size, the research was constrained by the number of cases due to strict inclusion and exclusion criteria, such as the exclusion of secondary and syndromic HCM cases. Furthermore, the investigation lacks a follow-up component for patients, which would have allowed for the assessment of long-term genotype-phenotype associations of specific genes in the development of HCM.

## Author Contributions

The study was conceptualized, designed and supervised by DSP. The clinical samples were provided by AR, KSM, HA, and JS. The data was analyzed by VJR, and TS. VJR and DSP drafted the manuscript. DSP reviewed the manuscript.

## Data Availability

The data that support the findings of this study are available from the corresponding author upon reasonable request.

## Abbreviations and Acronyms

HCM: Hypertrophic cardiomyopathy
SAI: South Asian Indian
P/LP: Pathogenic or likely pathogenic
VUS: Variant of uncertain significance
ECG: Electrocardiogram
LGE: Late gadolinium enhancement
ACMG: American College of Medical Genetics
LVIDd: Left ventricular internal diameter at diastole
LVIDs: Left ventricular internal diameter at end-systole
EF: Ejection fraction

