## Supplemental File for "Clinically actionable hypertrophic cardiomyopathy genes in South Asian Indians"

#### Supplemental Methods:

The patients were diagnosed as primary HCM using the following criteria:

1) Left ventricular hypertrophy ≥15 mm as observed on two-dimensional echocardiography, without any other underlying illness that accounts for the hypertrophy. Left ventricular septal to posterior wall thickness ratio greater than 1.3, without the presence of hypertension as a possible cause of hypertrophy.

2) Nonobstructive HCM was characterized by a pressure gradient of ≤30 mmHg at rest and after provocation. Patients with a pressure gradient >30 mmHg at rest or after provocation were categorized as having obstructive HCM.

3) Electrocardiogram (ECG) abnormalities that were present in the above echo-positive individuals include left ventricular hypertrophy (Romhilt-Estes score ≥4), Q-waves (with a duration >0.04 s and/or a depth >1/4 of ensuring R wave in at least two leads), and significant repolarization abnormalities (T-wave inversion in at least two leads). Non-sustained ventricular tachycardia was characterized by the occurrence of three or more ventricular extra systoles at a rate of ≥120 beats per minute, with a duration of <30 s.

Eighty-two individuals (24.48%) underwent cardiac magnetic resonance imaging (Cine, SSFP, T1-weighted, and T2-weighted imaging) using standard protocols. These individuals then underwent gadolinium administration and post-gadolinium imaging for late gadolinium enhancement (LGE) using a 1.5-T clinical system (Ingenia, Philips Healthcare, Best, The Netherlands). For LGE imaging, patients were given an intravenous injection of 0.2 mmol/kg of body weight gadolinium-based contrast reagent at a rate of 1 to 2 ml/s. After a 10-minute injection time, LGE images were acquired in the continuous short-axis view using an inversion-recovery gradient-recalled echo sequence with a manually selected optimal inversion time to minimize the signal from the normal myocardium.

##### Inclusion and exclusion criteria for primary HCM patients

- HCM patients, without any secondary or systemic diseases at the time of diagnosis were included
- Cases of HCM linked to conditions such as diabetes, hypertension, hyperlipidemia, coronary artery diseases, valvular diseases, and chronic renal failure, as well as those associated with smoking and alcohol use, were classified as "secondary HCM" and excluded from the study.

Additionally, the research did not include other forms of cardiomyopathy (such as dilated cardiomyopathy, left ventricular non-compaction, and arrhythmogenic right ventricular dysplasia), syndromic HCM cases, or pediatric HCM patients.

**Supplemental Table 1: List of clinically actionable genes categorized according to their association to HCM**

| Sl. No. | Gene Symbol | Protein Name |
|---|---|---|
| *Definitive* | | |
| 1 | *MYH7* | myosin heavy chain 7 |
| 2 | *MYBPC3* | myosin binding protein C3 |
| 3 | *TNNI3* | troponin I3 |
| 4 | *TNNT2* | troponin T2 |
| 5 | *TPM1* | tropomyosin 1 |
| 6 | *ACTC1* | actin alpha cardiac muscle 1 |
| 7 | *MYL2* | myosin light chain 2 |
| 8 | *MYL3* | myosin light chain 3 |
| 9 | *PRKAG2* | protein kinase AMP-activated non-catalytic subunit gamma 2 |
| 10 | *CSRP3* | cysteine and glycine rich protein 3 |
| 11 | *TNNC1* | troponin C1 |
| 12 | *FHOD3* | formin homology 2 domain containing 3 |
| *Strong* | | |
| 13 | *ALPK3* | alpha kinase 3 |
| *Moderate* | | |
| 14 | *JPH2* | junctophilin 2 |
| 15 | *TRIM63* | tripartite motif containing 63 |
| 16 | *KLHL24* | kelch like family member 24 |
| 17 | *MT-TI* | mitochondrially encoded tRNA isoleucine |
| *Limited* | | |
| 18 | *NEXN* | nexilin F-actin binding protein |
| 19 | *RPS6KB1* | ribosomal protein S6 kinase B1 |
| 20 | *RBM20* | RNA binding motif protein 20 |
| 21 | *OBSCN* | obscurin |
| 22 | *PDLIM3* | PDZ and LIM domain 3 |
| 23 | *KLF10* | KLF transcription factor 10 |
| 24 | *RYR2* | ryanodine receptor 2 |
| 25 | *TTN* | titin |
| 26 | *TMPO* | thymopoietin |
| *Disputed* | | |
| 27 | *MYLK2* | myosin light chain kinase 2 |
| 28 | *MYH6* | myosin heavy chain 6 |
| 29 | *KCNQ1* | potassium voltage-gated channel subfamily Q member 1 |
| 30 | *CASQ2* | calsequestrin 2 |
| 31 | *CALR3* | calreticulin 3 |
| 32 | *DSP* | desmoplakin |
| 33 | *MYOM1* | myomesin 1 |
| 34 | *TCAP* | titin-cap |
| 35 | *CACNB2* | calcium voltage-gated channel auxiliary subunit beta 2 |
| 36 | *MYOZ2* | myozenin 2 |
| 37 | *ANKRD1* | ankyrin repeat domain 1 |
| 38 | *VCL* | vinculin |
| 39 | *MYPN* | myopalladin |

#### Supplemental Table 2: Samples studied for SAI-HCM, SA Controls, other global HCM cohorts, and global controls

| Cohort name | Ethnicity | Number of samples | Reference | Clinically actionable genes analyzed |
|---|---|---|---|---|
| *Present Study* | | | | |
| SAI-HCM | South Indians | 335 | | All |
| **SA Controls** | | | | |
| gnomAD | South Asians | 45456 | https://gnomad.broadinstitute.org/ | All |
| Genome Asia 100K | South Asian Indians | 598 | GenomeAsia100K Consortium (31) | All |
| IndiGenomes | South Asian Indians | 1029 | Jain, A et al., (26) | All |
| i-DHANS | South Asian Indians | 94 | Dhandapany, PS et al., Jain, PK et al., (25,26) | All |
| **Other global HCM cohorts** | | | | |
| Europeans | Northern Europeans | 240 | Thompson, KL et al., (32) | *MYH6, MYLK2, NEXN, MYPN, TTN, KLF10, VCL, TCAP, CASQ2, MYOZ2, MYOM1, CALR3, KCNQ1* |
| Europeans | UK British | 505 | Lopes, LR et al., (33) | *PDLIM3* |
| North American | White | 302 | Chen, SN et al., (34) | *TRIM63* |
| Europeans | Europeans | 4867 | Salazar-Mendiguchia, J et al., (35) | *TRIM63* |
| Europeans | French | 5000 | Vadrot, N et al.,(36) | *TMPO* |
| European, South American and East Asians | Spanish, UK, Danes, Russians, Latvians, Brazilians, and Argentines, Chinese | 2817 | Lopes, LR et al., (18), Wang J et al., (37) | *ALPK3* |
| European and South American | Spanish, UK, United States, Denmark, Germany, and Argentina | 3189 | Ochoa, JP et al., (19) | *FHOD3* |
| UK | UK British | 684 | Allouba, M et al., (6) | *MYBPC3, MYH7, TNNT2, TNNI3, TPM1, MYL2, MYL3, CSRP3, ACTC1, TNNC1* |
| Middle Eastern | Egyptians | 514 | Allouba, M et al., (6) | |
| UK | UK British | 190 | Jain, PK et al., (26) | *RPS6KB1* |
| North Americans/United Kingdom | White | 2167-6179 | Pua, CJ et al., (20), Walsh R et al., (30) | *MYBPC3, MYH7, TNNT2, TNNI3, TPM1, MYL2, MYL3, CSRP3, ACTC1, TNNC1* |
| Southeast Asian | Singaporean | 224 | Pua, CJ et al., (20) | |
| East Asian | Chinese | 74 | Xu, J et al., (38) | *OBSCN* |
| **Global controls** | | | | |
| gnomAD | Global | 807162 | https://gnomad.broadinstitute.org/ | All |

#### Supplemental Table 3: Overall frequencies of case excess variants in clinically actionable moderate and limited gene categories in SAI-HCM

| Gene | cDNA Variant | Protein Variant | Variant Type | SAI-HCM frequency, n (%) | gnomad global frequency | gnomad SA frequency | ClinVar Classification | ACMG/AMP Classification | Final Classification |
|---|---|---|---|---|---|---|---|---|---|
| *Moderate* | | | | | | | | | |
| ***JPH2*** | c.1665_1666insTCG | p.S555_R556insS | ins | 1 (0.3) | 1.40E-06 | 2.04E-05 | NR | VUS | VUS |
| ***JPH2*** | c.G596A | p.G199D | mis | 1 (0.3) | 0 | 0 | NR | VUS | VUS |
| ***KLHL24*** | c.G724T | p.D242Y | mis | 1 (0.003) | 0 | 0 | NR | VUS | VUS |
| ***KLHL24*** | c.C517T | p.R173C | mis | 1 (0.003) | 0 | 0 | NR | VUS | VUS |
| *Limited* | | | | | | | | | |
| ***NEXN*** | c.851_864del | p.A284Gfs*2 | non | 1 (0.3) | 0 | 0 | NR | LP | LP |
| ***NEXN*** | c.C1171G | p.R391G | mis | 1 (0.3) | 6.847E-07 | 0 | NR | VUS | VUS |
| ***NEXN*** | c.C163A | p.Q55K | mis | 1 (0.3) | 6.85E-07 | 0 | NR | VUS | VUS |
| ***NEXN*** | c.C1523T | p.T508I | mis | 1 (0.3) | 0 | 0 | NR | VUS | VUS |
| ***NEXN*** | c.T1065G | p.D355E | mis | 1 (0.3) | 0 | 0 | NR | VUS | VUS |
| ***NEXN*** | c.G951A | p.M317I | mis | 1 (0.3) | 0 | 0 | NR | VUS | VUS |
| ***RPS6KB1*** | c.110delA | p. D37Afs*13 | non | 1 (0.003) | 0 | 0 | NR | LP | LP |
| ***RPS6KB1*** | c.C872T | p. P291L | mis | 1 (0.003) | 0 | 0 | NR | VUS | VUS |
| ***RPS6KB1*** | c.C1264G | p. P442A | mis | 1 (0.003) | 0 | 0 | NR | VUS | VUS |
| ***OBSCN*** | c.G18958A | p.D6320N | mis | 2 (0.006) | 0 | 0 | NR | VUS | VUS |
| ***OBSCN*** | c.G25561T | p.D8521Y | mis | 1 (0.003) | 0 | 0 | NR | VUS | VUS |
| ***OBSCN*** | c.A24005G | p.E8002G | mis | 2 (0.006) | 6.21E-07 | 0 | NR | VUS | VUS |
| ***OBSCN*** | c.G25162A | p.G8388S | mis | 2 (0.006) | 6.56E-07 | 0 | NR | VUS | VUS |
| ***OBSCN*** | c.25398_25402del | p.I8468Cfs*17 | non | 1 (0.003) | 0 | 0 | NR | NR | VUS |
| ***OBSCN*** | c.C23488A | p.L7830M | mis | 1 (0.003) | 0 | 0 | NR | VUS | VUS |
| ***OBSCN*** | c.10373delC | p.P3459Rfs*34 | non | 1 (0.003) | 0 | 0 | NR | NR | VUS |
| ***OBSCN*** | c.2240delC | p.P748Qfs*163 | non | 1 (0.003) | 0 | 0 | NR | NR | VUS |
| ***OBSCN*** | c.C17407G | p.Q5803E | mis | 1 (0.003) | 0 | 0 | NR | VUS | VUS |
| ***OBSCN*** | c.C20149T | p.Q6717X | mis | 1 (0.003) | 0 | 0 | NR | LP | LP |
| ***OBSCN*** | c.G6713A | p.R2238H | mis | 2 (0.006) | 4.96E-06 | 2.20E-05 | VUS | VUS | VUS |
| ***OBSCN*** | c.C16396T | p.R5466C | mis | 2 (0.006) | 1.37E-05 | 9.54E-05 | NR | VUS | VUS |
| ***OBSCN*** | c.G26483A | p.R8828H | mis | 1 (0.003) | 0 | 0 | NR | VUS | VUS |
| ***OBSCN*** | c.C6965A | p.S2322X | non | 1 (0.003) | 0 | 0 | NR | P | P |
| ***OBSCN*** | c.9578delC | p.S3194Afs*96 | non | 1 (0.003) | 0 | 0 | NR | NR | VUS |
| ***OBSCN*** | c.G25385A | p.S8462N | mis | 1 (0.003) | 6.20E-07 | 0 | NR | VUS | VUS |
| ***OBSCN*** | c.11945delC | p.T3983Qfs*8 | non | 1 (0.003) | 0 | 0 | NR | NR | VUS |
| ***OBSCN*** | c.C16568A | p.T5523K | mis | 1 (0.003) | 0 | 0 | NR | VUS | VUS |
| ***OBSCN*** | c.18711_18712insAC | p.T6240_T6241insI | ins | 1 (0.003) | 0 | 0 | NR | NR | VUS |
| ***OBSCN*** | c.C26432T | p.T8811I | mis | 1 (0.003) | 0 | 0 | NR | NR | VUS |
| ***RYR2*** | c.G14494A | p.E4832K | mis | 2 (0.006) | 3.10E-06 | 1.10E-05 | VUS | VUS | VUS |
| ***RYR2*** | c.T5156C | p.L1719P | mis | 1 (0.003) | 0 | 0 | NR | VUS | VUS |
| ***RYR2*** | c.C8968T | p.L2990F | mis | 1 (0.003) | 5.00E-06 | 1.11E-05 | VUS | VUS | VUS |
| ***RYR2*** | c.C9914G | p.P3305R | mis | 1 (0.003) | 0 | 0 | NR | VUS | VUS |
| ***RYR2*** | c.G9295A | p.V3099M | mis | 1 (0.003) | 0 | 0 | NR | VUS | VUS |
| ***RYR2*** | c.G152A | p.V718I | mis | 2 (0.006) | 0 | 0 | NR | VUS | VUS |
| ***TTN*** | c.C45092G | p.A15031G | mis | 1 (0.003) | 6.21E-07 | 1.10E-05 | NR | VUS | VUS |
| ***TTN*** | c.G50401T | p.A16801S | mis | 1 (0.003) | 0 | 0 | NR | VUS | VUS |
| ***TTN*** | c.C68210T | p.A22737V | mis | 1 (0.003) | 2.36E-05 | 2.21E-05 | NR | VUS | VUS |
| ***TTN*** | c.G88828A | p.A29610T | mis | 1 (0.003) | 1.24E-06 | 2.20E-05 | NR | VUS | VUS |
| ***TTN*** | c.32385_32838del | p.E10946Kfs*18 | non | 1 (0.003) | 0 | 0 | NR | NR | VUS |
| ***TTN*** | c.G34648A | p.E11550K | mis | 1 (0.003) | 0 | 0 | NR | VUS | VUS |
| ***TTN*** | c.G10639T | p.E3547X | non | 1 (0.003) | 0 | 0 | NR | VUS | VUS |
| ***TTN*** | c.14815delG | p.E4939Rfs*5 | non | 1 (0.003) | 0 | 0 | NR | VUS | VUS |
| ***TTN*** | c.82893_82894insCT | p.F27631_T27632i | ins | 2 (0.006) | 0 | 0 | NR | VUS | VUS |
| ***TTN*** | c.T90890C | p.F30297S | mis | 2 (0.006) | 0 | 0 | NR | VUS | VUS |
| ***TTN*** | c.G60673A | p.G20225R | mis | 1 (0.003) | 1.36E-05 | 1.10E-05 | VUS | VUS | VUS |
| ***TTN*** | c.G19264A | p.G6422R | mis | 1 (0.003) | 2.36E-05 | 0 | VUS | VUS | VUS |
| ***TTN*** | c.C60703T | p.H20235Y | mis | 1 (0.003) | 0 | 0 | NR | VUS | VUS |
| ***TTN*** | c.T40412C | p.I13471T | mis | 1 (0.003) | 6.40E-07 | 1.22E-05 | NR | VUS | VUS |
| ***TTN*** | c.A25508C | p.K8503T | mis | 1 (0.003) | 6.20E-07 | 1.10E-05 | NR | VUS | VUS |
| ***TTN*** | c.C98095T | p.L32699F | mis | 1 (0.003) | 5.63E-06 | 1.13E-05 | VUS | VUS | VUS |
| ***TTN*** | c.T96446C | p.M32149T | mis | 1 (0.003) | 1.86E-06 | 0 | NR | VUS | VUS |
| ***TTN*** | c.C62095G | p.P20699A | mis | 1 (0.003) | 0 | 0 | NR | VUS | VUS |
| ***TTN*** | c.12959delA | p.Q4320Rfs*25 | non | 1 (0.003) | 6.20E-07 | 0 | NR | NR | VUS |
| ***TTN*** | c.G15946T | p.A5316S | mis | 1 (0.003) | 0 | 0 | NR | VUS | VUS |
| ***TTN*** | c.C82933T | p.R27645C | mis | 1 (0.003) | 1.36E-05 | 2.20E-05 | Conflicting | LB | VUS |
| ***TTN*** | c.G85769T | p.R28590L | mis | 1 (0.003) | 1.86E-06 | 0 | NR | VUS | VUS |
| ***TTN*** | c.C89710T | p.R29904C | mis | 2 (0.006) | 3.72E-06 | 1.10E-05 | Conflicting | VUS | VUS |
| ***TTN*** | c.C92905T | p.R30969W | mis | 1 (0.003) | 1.55E-05 | 1.10E-05 | VUS | VUS | VUS |
| ***TTN*** | c.G99311A | p.R33104H | mis | 1 (0.003) | 6.83E-06 | 2.20E-05 | NR | VUS | VUS |
| ***TTN*** | c.A53674C | p.S17892R | mis | 1 (0.003) | 1.86E-06 | 1.10E-05 | VUS | VUS | VUS |
| ***TTN*** | c.C66503T | p.T22168I | mis | 1 (0.003) | 0 | 0 | NR | VUS | VUS |
| ***TTN*** | c.C17237T | p.T5746I | mis | 1 (0.003) | 0 | 0 | NR | VUS | VUS |
| ***TTN*** | c.G66847A | p.V22283I | mis | 1 (0.003) | 1.43E-05 | 2.20E-04 | VUS | VUS | VUS |
| ***TTN*** | c.82896dupA | p.V27633Sfs*5 | non | 2 (0.006) | 0 | 0 | NR | LP | LP |
| ***TTN*** | c.T10898G | p.V3633G | mis | 1 (0.003) | 1.86E-06 | 2.20E-05 | NR | VUS | VUS |
| ***TTN*** | c.T23875C | p.Y7959H | mis | 1 (0.003) | 6.20E-07 | 0 | NR | VUS | VUS |

**mis = missense; non = nonsense; ins = insertion; del = deletion; NR = not reported**

### Supplemental Table 4: Compound heterozygous, biallelic and multiallelic variants

| Compound heterozygous variants | | | | |
|---|---|---|---|---|
| 1 | ***MYBPC3*** | c.2869dupA | p.T957Nfs*93 | non |
| | ***MYH7*** | c.2623_2640del | p.E875_L880del | del |
| 2 | ***TNNT2*** | c.411_412insAAAGAGGA | p.I137_E138insKEEEELVSL | ins |
| | ***TPM1*** | c.226delA | p.K77Rfs*8 | non |
| 3 | ***MYH7*** | c.G1988A | p.R663H | mis |
| | ***MYH6*** | c.5046dupG | p.L1683Afs*3 | non |
| 4 | ***CSRP3*** | c.A224T | p.Q75L | mis |
| | ***KLHL24*** | c.G724T | p.D242Y | mis |
| 5 | ***DSP*** | c.C2697G | p.C899W | mis |
| | ***MYBPC3*** | c.C3372A | p.C1124X | non |
| 6 | ***DSP*** | c.7683_7685del | p.S2562del | del |
| | ***MYBPC3*** | c.C3811T | p.R1271X | non |
| 7 | ***FHOD3*** | c.G1040A | p.C347Y | mis |
| | ***MYH7*** | c.C1357T | p.R453C | mis |
| | ***RBM20*** | c.A995T | p.D332V | mis |
| 8 | ***OBSCN*** | c.A24005G | p.E8002G | mis |
| | ***MYBPC3*** | c.G1730A | p.W577X | non |
| 9 | ***MYL2*** | c.G64A | p.E22K | mis |
| | ***VCL*** | c.G563A | p.R188Q | mis |
| 10 | ***MYH7*** | c.T2207C | p.I736T | mis |
| | ***MYOM1*** | c.T3331G | p.Y1111D | mis |
| 11 | ***KLHL24*** | c.C517T | p.R173C | mis |
| | ***TTN*** | c.G34648A | p.E11550K | mis |
| 12 | ***MYH6*** | c.G4446C | p.E1482D | mis |
| | ***TTN*** | c.T10898G | p.V3633G | mis |
| 13 | ***MYBPC3*** | c.3746delG | p.G1249Afs*82 | non |
| | ***TTN*** | c.C98095T | p.L32699F | mis |
| 14 | ***ALPK3*** | c.635_636del | p.L213Afs*30 | non |
| | ***TMPO*** | c.A1901C | p.D634A | mis |
| 15 | ***MYH7*** | c.G1816A | p.V606M | mis |
| | ***OBSCN*** | c.C26432T | p.T8811I | mis |
| 16 | ***MYH7*** | c.G1816A | p.V606M | mis |
| | ***OBSCN*** | c.G26483A | p.R8828H | mis |
| | ***OBSCN*** | c.C6965A | p.S2322X | non |
| 17 | ***MYBPC3*** | c.G1171T | p.D391Y | mis |
| | ***MYBPC3*** | c.989delC | p.P330Hfs*20 | non |
| | ***TTN*** | c.32385_32838del | p.E10946Kfs*18 | non |
| 18 | ***MYBPC3*** | c.C3372A | p.C1124X | non |
| | ***OBSCN*** | c.G6713A | p.R2238H | mis |
| 19 | ***MYBPC3*** | c.C1112T | p.P371L | mis |
| | ***TTN*** | c.C68210T | p.A22737V | mis |
| 20 | ***KCNQ1*** | c.G1006T | p.A336S | mis |
| | ***MYBPC3*** | c.C1358T | p.P453Lfs*13 | non |

| | | | | |
|---|---|---|---|---|
| 21 | ***MYH7*** | c.C4258T | p.R1420W | mis |
| | ***TTN*** | c.12959delA | p.Q4320Rfs*25 | non |
| 22 | ***ALPK3*** | c.G2374T | p.G792C | mis |
| | ***NEXN*** | c.851_864del | p.A284Gfs*3 | non |
| 23 | ***MYBPC3*** | c.G2459A | p.R820Q | mis |
| | ***OBSCN*** | c.G25385A | p.S8462N | mis |
| 24 | ***PRKAG2*** | c.1053_1067del | p.L352_E356del | del |
| | ***TTN*** | c.T96446C | p.M32149T | mis |
| 25 | ***JPH2*** | c.G596A | p.G199D | mis |
| | ***OBSCN*** | c.2240delC | p.P748Qfs*163 | non |
| 26 | ***MYH6*** | c.G2334C | p.M778I | mis |
| | ***OBSCN*** | c.11945delC | p.T3983Qfs*8 | non |
| 27 | ***CSRP3*** | c.A155C | p.H52P | mis |
| | ***TTN*** | c.C82933T | p.R27645C | mis |
| 28 | ***MYH6*** | c.A5573G | p.E1858G | mis |
| | ***TTN*** | c.A25508C | p.K8503T | mis |
| 29 | ***FHOD3*** | c.A2239T | p.S747C | mis |
| | ***MYH6*** | c.A5573G | p.E1858G | mis |
| 30 | ***MYH6*** | c.A5573G | p.E1858G | mis |
| | ***MYOM1*** | c.G3200A | p.R1067Q | mis |
| 31 | ***MYH6*** | c.C1720T | p.Q574X | non |
| | ***MYBPC3*** | c.T620C | p.L207P | mis |
| | ***TTN*** | c.G15946T | p.A5316S | mis |
| | ***TTN*** | c.C60703T | p.H20235Y | mis |
| | ***TTN*** | c.G85769T | p.R28590L | mis |
| | ***TTN*** | c.G50401T | p.A16801S | mis |
| 32 | ***CACNB2*** | c.37_39del | p.A16del | del |
| | ***MYH6*** | c.A5573G | p.E1858G | mis |
| 33 | ***FHOD3*** | c.1197_1199del | p.E406del | del |
| | ***TMPO*** | c.241_243del | p.A84del | del |
| | ***TTN*** | c.G99311A | p.R33104H | mis |
| 34 | ***CACNB2*** | c.G1956T | p.W652C | mis |
| | ***OBSCN*** | c.C9268T | p.R3090C | mis |
| 35 | ***MYH6*** | c.A5573G | p.E1858G | mis |
| | ***OBSCN*** | c.C17407G | p.Q5803E | mis |
| | ***TTN*** | c.C66503T | p.T22168I | mis |

| Biallelic and multiallelic variants | | | | |
|---|---|---|---|---|
| 1 | ***DSP*** | c.A5963G | p.K1988R | mis |
| | ***DSP*** | c.G3806A | p.R1269Q | mis |
| 2 | ***TTN*** | c.T40412C | p.I13471T | mis |
| | ***TTN*** | c.G19264A | p.G6422R | mis |
| 3 | ***OBSCN*** | c.C16396T | p.R5466C | mis |
| | ***OBSCN*** | c.C23488A | p.L7830M | mis |
| 4 | ***NEXN*** | c.254dupA | p.D85Efs*5 | non |
| | ***NEXN*** | c.258del | p.S87Lfs*4 | non |
| 5 | ***TTN*** | c.82893_82894insCTTGTA | p.F27631_T27632insLVI | ins |
| | ***TTN*** | c.82896dupA | p.V27633Sfs*5 | non |
| 6 | ***TTN*** | c.82893_82894insCTTGTA | p.F27631_T27632insLVI | ins |
| | ***TTN*** | c.82896dupA | p.V27633Sfs*5 | non |
| | ***TTN*** | c.G60673A | p.G20225R | mis |

### Supplemental Table 5: Prevalence of P/LP/exVUS and case excess in clinically actionable HCM genes in SAI-HCM cohort

| Gene | SAI-HCM (n=335) | | | Other HCM (n= 74 - 7601)# | | | Rare* control VUS frequency, n (%) | | Fisher's exact p - value | | | | SAI-HCM exVUS† | | SAI-HCM total case excess$ | | Other HCM exVUS† | | Other HCM total case excess$ | |
|---|---|---|---|---|---|---|---|---|---|---|---|---|---|---|---|---|---|---|---|---|
| | P/LP frequency, n (%) | VUS frequency, n (%) | Total, n (%) | P/LP frequency, n (%) | VUS frequency, n (%) | Total, n (%) | Global (gnomAD, n=807162) | SA (gnomAD, IndiGenomes, GenomeAsia and iDhans, n=47117) | Global control vs SAI-HCM | Global control vs Other HCM | SA control vs SAI-HCM | SA control vs Other HCM | Global % | SA % | Global % | SA % | Global % | SA % | Global % | SA % |
| *Definitive* | | | | | | | | | | | | | | | | | | | | |
| *MYBPC3* | 20 (5.97) | 2 (0.59) | 22 (6.56) | 1062 (13.97) | 322 (4.23) | 1384 (18.2) | 8091 (1) | 379 (0.79) | <0.0001 | <0.0001 | <0.0001 | <0.0001 | No excess | No excess | 5.56 | 5.77 | 3.23 | 3.44 | 17.2 | 17.41 |
| *MYH7* | 17 (5.07) | 7 (2.08) | 24 (7.15) | 744 (9.87) | 318 (4.22) | 1062 (14.09) | 7328 (0.90) | 355 (0.74) | <0.0001 | <0.0001 | <0.0001 | <0.0001 | 1.18 | 1.34 | 6.25 | 6.41 | 3.32 | 3.48 | 13.19 | 13.35 |
| *TNNT2* | 1 (0.29) | 3 (0.89) | 4 (1.15) | 82 (1.08) | 63 (0.83) | 145 (1.92) | 1237 (0.15) | 73 (0.15) | 0.0538 | <0.0001 | 0.0013 | <0.0001 | 0.74 | 0.74 | 1 | 1 | 0.68 | 0.68 | 1.77 | 1.77 |
| *TNNI3* | 0 | 0 | 0 | 105 (1.40) | 65 (0.87) | 170 (2.27) | 999 (0.12) | 68 (0.14) | 0.8988 | <0.0001 | 0.5848 | <0.0001 | 0 | 0 | 0 | 0 | 0.75 | 0.73 | 2.15 | 2.13 |
| *TPM1* | 1 (0.29) | 1 (0.29) | 2 (0.58) | 33 (0.56) | 46 (0.78) | 79 (1.34) | 693 (0.085) | 66 (0.13) | 0.2356 | <0.0001 | 0.0602 | <0.0001 | 0.20 | 0.16 | 0.49 | 0.45 | 0.695 | 0.65 | 1.255 | 1.21 |
| *ACTC1* | 0 | 0 | 0 | 8 (0.14) | 20 (0.35) | 28 (0.49) | 260(0.032) | 14 (0.029) | 0.57 | <0.0001 | 0.7525 | <0.0001 | 0 | 0 | 0 | 0 | 0.318 | 0.321 | 0.458 | 0.461 |
| *MYL2* | 1 (0.29) | 2 (0.59) | 3 (0.88) | 27 (0.48) | 35 (0.62) | 62 (1.1) | 949 (0.117) | 62 (0.13) | 0.0108 | <0.0001 | 0.0012 | <0.0001 | 0.47 | 0.46 | 0.76 | 0.75 | 0.503 | 0.49 | 0.983 | 0.97 |
| *MYL3* | 0 | 0 | 0 | 10 (0.17) | 46 (0.82) | 56 (0.99) | 958 (0.119) | 36 (0.076) | 0.972 | <0.0001 | 0.5985 | <0.0001 | 0 | 0 | 0 | 0 | 0.701 | 0.744 | 0.871 | 0.914 |
| *PRKAG2* | 0 | 3 (0.89) | 3 (0.89) | 12 (0.28) | 30 (0.71) | 42 (1) | 1098 (0.24) | 106 (0.22) | 0.7277 | <0.0001 | 0.315 | <0.0001 | 0.65 | 0.67 | 0.65 | 0.607 | 0.47 | 0.49 | 0.76 | 0.78 |
| *CSRP3* | 1 (0.29) | 1 (0.29) | 2 (0.58) | 8 (0.22) | 23 (0.64) | 31 (0.86) | 1419 (0.18) | 69 (0.14) | 0.8274 | <0.0001 | 0.6527 | <0.0001 | 0.11 | 0.15 | 0.40 | 0.44 | 0.46 | 0.5 | 0.68 | 0.72 |
| *TNNC1* | 0 | 0 | 0 | 0 | 5 (0.1) | 5 (0.1) | 417 (0.05) | 25 (0.05) | 0.7689 | 0.3107 | 0.686 | 0.8404 | 0 | 0 | 0 | 0 | 0.05 | 0.05 | 0.05 | 0.05 |
| *FHOD3* | 2 (0.59) | 4 (1.19) | 6 (1.78) | 35 (1.09) | 39 (0.80) | 74 (1.89) | 519 (0.064) | 382 (0.8) | <0.0001 | <0.0001 | <0.0001 | <0.0001 | 1.13 | 0.39 | 1.716 | 0.98 | 0.736 | 0 | 1.826 | 1.09 |
| *Strong* | | | | | | | | | | | | | | | | | | | | |
| *ALPK3* | 6 (1.79) | 2 (0.59) | 8 (2.38) | 39 (1.38) | 5 (0.17) | 44 (1.56) | 7775 (0.96) | 277 (0.58) | 0.0005 | <0.0001 | <0.0001 | <0.0001 | No excess | 0.01 | 1.42 | 1.8 | No excess | No excess | 0.6 | 0.98 |
| *Moderate* | | | | | | | | | | | | | | | | | | | | |
| *JPH2* | 0 | 2 (0.59) | 2 (0.59) | 0 | 17 (1.41) | 17 (1.41) | 2854 (0.35) | 184 (0.38) | 0.856 | 0.0003 | 0.6933 | <0.0001 | 0.24 | 0.21 | 0.24 | 0.21 | 1.06 | 1.03 | 1.06 | 1.03 |
| *TRIM63* | 0 | 0 | 0 | 0 | 3 (0.06) | 3 (0.06) | 82 (0.01) | 10 (0.02) | 0.0115 | 0.0075 | 0.7798 | <0.0001 | 0.00 | 0.00 | 0.00 | 0.00 | 0.05 | 0.00 | 0.05 | 0.00 |
| *KLHL24* | 0 | 2 (0.60) | 2 (0.60) | Ins. Nos. | Ins. Nos. | Ins. Nos. | 295 (0.04) | 42 (0.08) | <0.0001 | NC | 0.032 | NC | 0.56 | 0.00 | 0.56 | 0.00 | 0.00 | 0.00 | 0.00 | 0.00 |
| *MT-TI* | 0 | 0 | 0 | Ins. Nos. | Ins. Nos. | Ins. Nos. | 0 | 0 | NC | NC | NC | NC | 0.00 | 0.00 | 0.00 | 0.00 | 0.00 | 0.00 | 0.00 | 0.00 |
| *Limited* | | | | | | | | | | | | | | | | | | | | |
| *NEXN* | 1 (0.29) | 5 (1.49) | 6 (1.78) | 0 | 1 (0.41) | 1 (0.41) | 2868 (0.36) | 176 (0.37) | 0.0096 | 0.6079 | <0.0001 | 0.4842 | 1.13 | 1.12 | 1.42 | 1.41 | 0.05 | 0.04 | 0.05 | 0.04. |
| *RBM20* | 0 | 3 (0.90) | 3 (0.90) | 1 (0.13) | 18 (2.27) | 19 (2.40) | 6637 (0.82) | 201 (0.41) | 0.8775 | <0.0001 | 0.3997 | <0.0001 | 0.00 | 0.59 | 0.08 | 0.59 | 1.45 | 1.86 | 1.45 | 1.86 |
| *RPS6KB1* | 1 (0.29) | 2 (0.59) | 3 (0.89) | 2 (1.05) | 0 | 2 (1.05) | 30 (0.004) | 58 (0.12) | <0.0001 | <0.0001 | <0.0001 | <0.0001 | 0.586 | 0.47 | 0.886 | 0.77 | 0 | 0 | 1.046 | 0.93 |
| *OBSCN* | 2 (0.60) | 12 (3.58) | 14 (4.18) | 0 | 1 (1.35) | 1 (1.35) | 17093 (2.18) | 718 (1.42) | 0.0151 | 0.9568 | 0.0003 | 0.8902 | 1.400 | 2.16 | 2 | 2.76 | No excess | No excess | No excess | No excess |
| *PDLIM3* | 0 | 0 | 0 | 0 | 1 (0.20) | 1 (0.20) | 1973 (0.24) | 66 (0.14) | 0.7244 | 0.8107 | 0.4868 | <0.0001 | 0.00 | 0.00 | 0.00 | 0.00 | No excess | 0.06 | No excess | 0.06 |
| *KLF10* | 0 | 0 | 0 | 0 | 1 (0.42) | 1 (0.42) | 767 (0.10) | 53 (0.10) | 0.7471 | 0.5693 | 0.5393 | 0.6637 | 0.00 | 0.00 | 0.00 | 0.00 | 0.32 | 0.32 | 0.32 | 0.32 |
| *RYR2* | 0 | 8 (2.39) | 8 (2.39) | 0 | 2 (2.70) | 2 (2.70) | 19123 (2.37) | 594 (1.28) | 0.8753 | 0.8465 | 0.132 | 0.5818 | 0.02 | 1.11 | 0.02 | 1.11 | 0.33 | 1.42 | 0.33 | 1.42 |
| *TTN* | 3 (0.90) | 31 (9.25) | 34 (10.15) | 0 | 87 (36.25) | 87 (36.25) | 122759 (15.21) | 4221 (8.46) | 0.0123 | <0.0001 | 0.6148 | <0.0001 | No excess | 0.79 | No excess | 1.69 | 21.04 | 27.79 | 21.04 | 27.79 |
| *TMPO* | 0 | 1 (0.30) | 1 (0.30) | 0 | 1 (0.02) | 1 (0.02) | 3126 (0.39) | 110 (0.22) | 0.8584 | <0.0001 | 0.8052 | 0.0032 | No excess | 0.08 | No excess | 0.08 | No excess | No excess | No excess | No excess |
| *Disputed* | | | | | | | | | | | | | | | | | | | | |
| *MYLK2* | 1 (0.29) | 0 | 1 (0.29) | 0 | 0 | 0 | 2241 (0.28) | 133 (0.28) | 0.7501 | 0.7171 | 0.5512 | 0.5284 | 0 | 0 | 0.01 | 0.01 | 0 | 0 | 0 | 0 |
| *MYH6* | 3 (0.89) | 18 (5.37) | 21 (6.26) | 0 | 7 (2.91) | 7 (2.91) | 9737 (1.2) | 434 (0.91) | <0.0001 | 0.9594 | <0.0001 | 0.2613 | 4.17 | 4.46 | 5.06 | 5.35 | 1.71 | 2 | 1.71 | 2 |
| *KCNQ1* | 0 | 1 (0.30) | 1 (0.30) | 0 | 6 (2.5) | 6 (2.5) | 4558 (0.56) | 151 (0.32) | 0.7753 | 0.0004 | 0.9287 | <0.0001 | No excess | No excess | No excess | No excess | 1.94 | 1.94 | 2.18 | 2.18 |
| *CASQ2* | 0 | 1 (0.30) | 1 (0.30) | 0 | 1 (0.42) | 1 (0.42) | 1997 (0.25) | 54 (0.12) | 0.7175 | 0.9029 | 0.8667 | 0.6833 | 0.25 | 0.18 | 0.25 | 0.18 | 0.17 | 0.30 | 0.17 | 0.30 |
| *CALR3* | 0 | 0 | 0 | 0 | 2 (0.83) | 2 (0.83) | 1375 (0.18) | 39 (0.08) | 0.9256 | 0.0879 | 0.5939 | 0.0051 | 0.00 | 0.00 | 0.00 | 0.00 | 0.65 | 0.75 | 0.65 | 0.75 |
| *DSP* | 1 (0.30) | 6 (1.79) | 7 (2.09) | 0 | 7 (0.70) | 7 (0.70) | 14991 (1.86) | 405 (2.09) | 0.9104 | 0.0095 | 0.0401 | 0.6606 | No excess | No excess | No excess | No excess | No excess | No excess | No excess | No excess |
| *MYOM1* | 0 | 2 (0.60) | 2 (0.60) | 0 | 5 (2.08) | 5 (2.08) | 8449 (1.05) | 293 (0.60) | 0.589 | 0.2076 | 0.9138 | 0.0182 | No excess | No excess | No excess | No excess | 1.03 | 1.48 | 1.03 | 1.48 |
| *TCAP* | 0 | 0 | 0 | 0 | 1 (0.42) | 1 (0.42) | 1194 (0.15) | 41 (0.09) | 0.9947 | 0.8079 | 0.5893 | 0.5319 | 0.00 | 0.00 | 0.00 | 0.00 | 0.27 | 0.33 | 0.27 | 0.33 |
| *CACNB2* | 1 (0.30) | 1 (0.30) | 2 (0.60) | Ins. Nos. | Ins. Nos. | Ins. Nos. | 3125 (.39) | 103 (0.22) | 0.8584 | NC | 0.402 | NC | No excess | 0.08 | 0.41 | 0.38 | 0.00 | 0.00 | 0.00 | 0.00 |
| *MYOZ2* | 0 | 0 | 0 | 0 | 1 (0.42) | 1 (0.42) | 943 (0.12) | 27 (0.06) | 0.8618 | 0.6785 | 0.6555 | 0.3553 | 0.00 | 0.00 | 0.00 | 0.00 | 0.30 | 0.36 | 0.30 | 0.36 |
| *ANKRD1* | 0 | 1 (0.30) | 1 (0.30) | Ins. Nos. | Ins. Nos. | Ins. Nos. | 1551 (0.19) | 74 (0.14) | 0.8575 | NC | 0.5423 | NC | 0.11 | 0.16 | 0.11 | 0.16 | 0.00 | 0.00 | 0.00 | 0.00 |
| *VCL* | 0 | 1 (0.30) | 1 (0.30) | 0 | 1 (0.42) | 1 (0.42) | 4749 (0.59) | 138 (0.30) | 0.7374 | 0.9399 | 0.9877 | 0.75 | No excess | No excess | No excess | No excess | No excess | 0.12 | No excess | 0.12 |
| *MYPN* | 0 | 0 | 0 | 0 | 3 (1.25) | 3 (1.25) | 5091 (0.63) | 294 (0.62) | 0.2209 | 0.543 | 0.4923 | 0.1006 | 0 | 0 | 0 | 0 | 0.62 | 0.63 | 0.62 | 0.63 |

P, pathogenic; LP, likely pathogenic; SA, South Asian; Other HCM, other global HCM case; exVUS, excess of variant variants of unknown significance;

NC, Not comparable; Ins. Nos., Insignificant numbers; cannot be used for analysis; #, Sample range breakdown detailed in Supplementary Table 2; Zero (0) indicates no variants identified; No Excess indicates the prevalence of variants in

**Supplemental Table 6: The status of P/LP variants in other 166 inheritable cardiac disease genes in SAI-HCM**

| Other known cardiac disease gene | Variant Present/ Absent | Other known cardiac disease gene | Variant Present/ Absent | Other known cardiac disease gene | Variant Present/ Absent | Other known cardiac disease gene | Variant Present/ Absent |
|---|---|---|---|---|---|---|---|
| *A2ML1* | *Absent* | *CTF1* | *Absent* | *KCNH2* | *Absent* | *RIT1* | *Absent* |
| *ABCC9* | *Absent* | *DES* | *Absent* | *KCNJ2* | *Absent* | *RYR1* | *Absent* |
| *ABCG5* | *Absent* | *DMD* | *Absent* | *KCNJ5* | *Absent* | *SALL4* | *Absent* |
| *ABCG8* | *Absent* | *DNAJC19* | *Absent* | *KCNJ8* | *Absent* | *SCN1B* | *Absent* |
| *ABL1* | *Absent* | *DOLK* | *Absent* | *KRAS* | *Absent* | *SCN2B* | *Absent* |
| *ACADVL* | *Absent* | *DPP6* | *Absent* | *LAMA2* | *Absent* | *SCN3B* | *Absent* |
| *ACTA1* | *Absent* | *DSC2* | *Absent* | *LAMA4* | *Absent* | *SCN4B* | *Absent* |
| *ACTA2* | *Absent* | *DSG2* | *Absent* | *LAMP2* | *Absent* | *SCN5A* | *Absent* |
| *ACTN2* | *Absent* | *DTNA* | *Absent* | *LDB3* | *Absent* | *SCO2* | *Absent* |
| *ACVR2B* | *Absent* | *EFEMP2* | *Absent* | *LDLR* | *Absent* | *SDHA* | *Absent* |
| *ACVRL1* | *Absent* | *ELN* | *Absent* | *LDLRAP1* | *Absent* | *SEPN1* | *Absent* |
| *ADA2* | *Absent* | *EMD* | *Absent* | *LMF1* | *Absent* | *SGCB* | *Absent* |
| *ADAMTS10* | *Absent* | *EYA4* | *Absent* | *LMNA* | *Absent* | *SGCD* | *Absent* |
| *ADAMTS17* | *Absent* | *FBN1* | *Absent* | *LPL* | *Absent* | *SGCG* | *Absent* |
| *AGL* | *Absent* | *FBN2* | *Absent* | *LTBP2* | *Absent* | *SHOC2* | *Absent* |
| *AKAP9* | *Absent* | *FHL1* | *Absent* | *LZTR1* | *Present (1)* | *SLC25A4* | *Absent* |
| *ALMS1* | *Absent* | *FHL2* | *Absent* | *MAP2K1* | *Absent* | *SLC2A10* | *Absent* |
| *ANK2* | *Absent* | *FKRP* | *Absent* | *MAP2K2* | *Absent* | *SMAD3* | *Absent* |
| *APOA4* | *Absent* | *FKTN* | *Absent* | *MIB1* | *Absent* | *SMAD4* | *Absent* |
| *APOA5* | *Absent* | *FXN* | *Absent* | *MRPL3* | *Absent* | *SNTA1* | *Absent* |
| *APOB* | *Absent* | *GAA* | *Absent* | *MTO1* | *Absent* | *SOS1* | *Absent* |
| *APOC2* | *Absent* | *GATAD1* | *Absent* | *MURC* | *Absent* | *SREBF2* | *Absent* |
| *APOE* | *Absent* | *GCKR* | *Absent* | *MYH11* | *Absent* | *SVIL* | *Absent* |
| *BAG3* | *Absent* | *GJA5* | *Absent* | *MYLK* | *Absent* | *TAZ* | *Absent* |
| *BRAF* | *Absent* | *GLA* | *Absent* | *MYO6* | *Absent* | *TBX20* | *Absent* |
| *CACNA1C* | *Absent* | *GPD1L* | *Absent* | *NKX2-5* | *Absent* | *TBX3* | *Absent* |
| *CACNA2D1* | *Absent* | *GPIHBP1* | *Absent* | *NODAL* | *Absent* | *TBX5* | *Absent* |
| *CALM1* | *Absent* | *GYG1* | *Absent* | *NOTCH1* | *Absent* | *TGFB2* | *Absent* |
| *CALR3* | *Absent* | *HADHA* | *Absent* | *NPHP3* | *Absent* | *TGFB3* | *Absent* |
| *CAV1* | *Absent* | *HAND1* | *Absent* | *NPPA* | *Absent* | *TGFBR1* | *Absent* |
| *CAV3* | *Absent* | *HCN4* | *Absent* | *NRAS* | *Absent* | *TGFBR2* | *Absent* |
| *CBL* | *Absent* | *HFE* | *Absent* | *PCSK9* | *Absent* | *TMEM43* | *Absent* |
| *CBS* | *Absent* | *HRAS* | *Absent* | *PKD1L1* | *Absent* | *TRDN* | *Absent* |
| *CETP* | *Absent* | *HSPB8* | *Absent* | *PKP2* | *Absent* | *TRPM4* | *Absent* |
| *CHRM2* | *Absent* | *ILK* | *Absent* | *PLN* | *Absent* | *TTC8* | *Absent* |
| *COL3A1* | *Absent* | *IPO8* | *Absent* | *PRDM16* | *Absent* | *TTR* | *Absent* |
| *COL5A1* | *Absent* | *JAG1* | *Absent* | *PRKAR1A* | *Absent* | *TXNRD2* | *Absent* |
| *COL5A2* | *Absent* | *JUP* | *Absent* | *PSEN1* | *Absent* | *ZBTB17* | *Absent* |
| *COX15* | *Absent* | *KCNA5* | *Absent* | *PSEN2* | *Absent* | *ZHX3* | *Absent* |
| *CREB3L3* | *Absent* | *KCNE1* | *Absent* | *PTPN11* | *Absent* | *ZIC3* | *Absent* |
| *CRELD1* | *Absent* | *KCNE2* | *Absent* | *RAF1* | *Absent* | | |
| *CRYAB* | *Absent* | *KCNE3* | *Absent* | *RANGRF* | *Absent* | | |

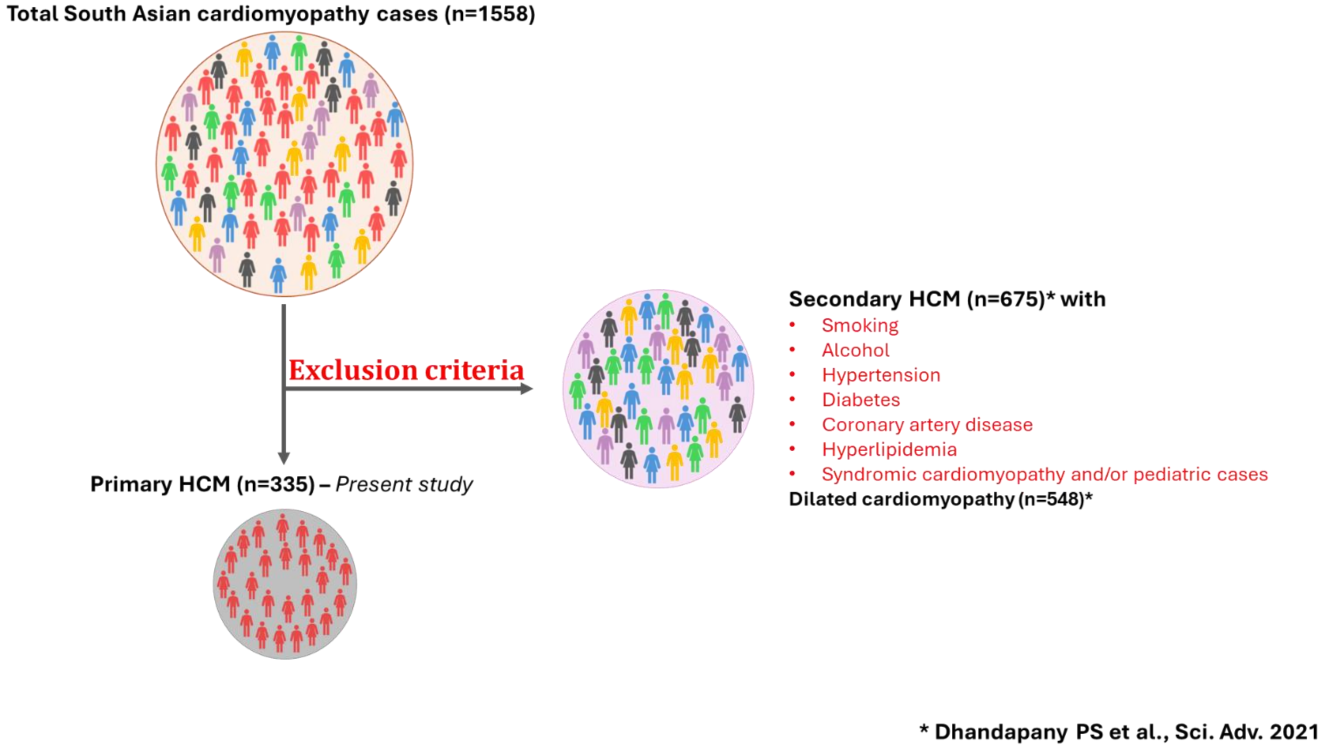

**Supplemental Figure 1: Primary SAI-HCM patient selection schematic. Three hundred thirty-five primary HCM cases were included for this study**

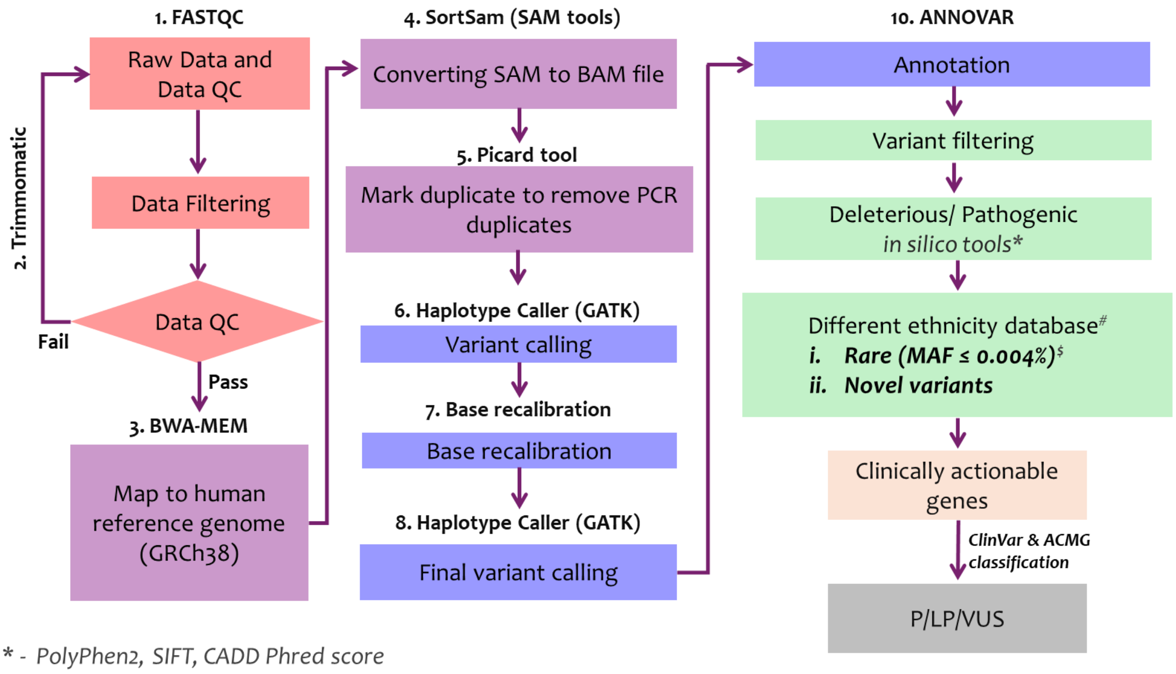

** - PolyPhen2, SIFT, CADD Phred score*
*# - Mixed control databases (gnomAD (v4.1), iDHANS, Genome Asia 100K, The IndiGenomes) and disease databases (Geno2MP v2.4, and ClinVar)*
$ - *Whiffin N* et al. *Genet Med (2019)*

**Supplemental Figure 2: Exome sequencing analysis pipeline to identify clinically actionable gene variants in SAI-HCM.**
